# Morphological, cellular and molecular basis of brain infection in COVID-19 patients

**DOI:** 10.1101/2020.10.09.20207464

**Authors:** Fernanda Crunfli, Victor Corasolla Carregari, Flavio Protasio Veras, Pedro Henrique Vendramini, Aline Gazzola Fragnani Valença, André Saraiva Leão Marcelo Antunes, Caroline Brandão-Teles, Giuliana da Silva Zuccoli, Guilherme Reis-de-Oliveira, Lícia C. Silva-Costa, Verônica Monteiro Saia-Cereda, Bradley Joseph Smith, Ana Campos Codo, Gabriela Fabiano de Souza, Stéfanie Primon Muraro, Pierina Lorencini Parise, Daniel A. Toledo-Teixeira, Ícaro Maia Santos de Castro, Bruno Marcel Silva Melo, Glaucia M. Almeida, Egidi Mayara Silva Firmino, Isadora Marques Paiva, Bruna Manuella Souza Silva, Rafaela Mano Guimarães, Niele D. Mendes, Raíssa Guimarães Ludwig, Gabriel Palermo Ruiz, Thiago Leite Knittel, Gustavo Gastão Davanzo, Jaqueline Aline Gerhardt, Patrícia Brito Rodrigues, Julia Forato, Mariene Ribeiro Amorim, Natália Brunetti Silva, Matheus Cavalheiro Martini, Maíra Nilson Benatti, Sabrina Batah, Li Siyuan, Rafael Batista João, Lucas Scardua Silva, Mateus Henrique Nogueira, Ítalo Karmann Aventurato, Mariana Rabelo de Brito, Marina Koutsodontis Machado Alvim, José Roberto da Silva Júnior, Lívia Liviane Damião, Iêda Maria Pereira de Sousa, Elessandra Dias da Rocha, Solange Maria Gonçalves, Luiz Henrique Lopes da Silva, Vanessa Bettini, Brunno Machado de Campos, Guilherme Ludwig, Lucas Alves Tavares, Marjorie Cornejo Pontelli, Rosa Maria Mendes Viana, Ronaldo Martins, Andre S. Vieira, José Carlos Alves-Filho, Eurico Arruda, Guilherme Podolski-Gondim, Marcelo Volpon Santos, Luciano Neder, Fernando Cendes, Paulo Louzada-Junior, Renê Donizeti Oliveira, Fernando Queiroz Cunha, André Damásio, Marco Aurélio Ramirez Vinolo, Carolina Demarchi Munhoz, Stevens K. Rehen, Helder I Nakaya, Thais Mauad, Amaro Nunes Duarte-Neto, Luiz Fernando Ferraz da Silva, Marisa Dolhnikoff, Paulo Saldiva, Alessandro S. Farias, Pedro Manoel M. Moraes-Vieira, Alexandre Todorovic Fabro, Adriano S. Sebollela, José Luiz Proença Módena, Clarissa Lin Yasuda, Marcelo A. Mori, Thiago Mattar Cunha, Daniel Martins-de-Souza

**Author notes:** Correspondence should be addressed to: Daniel Martins-de-Souza, PhD, Thiago Mattar Cunha, PhD, Marcelo A. Mori, PhD, Clarissa Lin Yasuda, MD, PhD. These authors contributed equally to this work (Fernanda Crunfli, Victor Corasolla Carregari, Flavio Protasio Veras).

## Abstract

Although increasing evidence confirms neuropsychiatric manifestations associated mainly with severe COVID-19 infection, the long-term neuropsychiatric dysfunction has been frequently observed after mild infection. Here we show the spectrum of the cerebral impact of SARS-CoV-2 infection ranging from long-term alterations in mildly infected individuals (orbitofrontal cortical atrophy, neurocognitive impairment, excessive fatigue and anxiety symptoms) to severe acute damage confirmed in brain tissue samples extracted from the orbitofrontal region (via endonasal trans-ethmoidal approach) from individuals who died of COVID-19. We used surface-based analyses of 3T MRI and identified orbitofrontal cortical atrophy in a group of 81 mildly infected patients (77% referred anosmia or dysgeusia during acute stage) compared to 145 healthy volunteers; this atrophy correlated with symptoms of anxiety and cognitive dysfunction. In an independent cohort of 26 individuals who died of COVID-19, we used histopathological signs of brain damage as a guide for possible SARS-CoV-2 brain infection, and found that among the 5 individuals who exhibited those signs, all of them had genetic material of the virus in the brain. Brain tissue samples from these 5 patients also exhibited foci of SARS-CoV-2 infection and replication, particularly in astrocytes. Supporting the hypothesis of astrocyte infection, neural stem cell-derived human astrocytes *in vitro* are susceptible to SARS-CoV-2 infection through a non-canonical mechanism that involves spike-NRP1 interaction. SARS-CoV-2-infected astrocytes manifested changes in energy metabolism and in key proteins and metabolites used to fuel neurons, as well as in the biogenesis of neurotransmitters. Moreover, human astrocyte infection elicits a secretory phenotype that reduces neuronal viability. Our data support the model in which SARS-CoV-2 reaches the brain, infects astrocytes and consequently leads to neuronal death or dysfunction. These deregulated processes are also likely to contribute to the structural and functional alterations seen in the brains of COVID-19 patients.

## INTRODUCTION

COVID-19 is a disease caused by infection with the severe acute respiratory syndrome coronavirus 2 (SARS-CoV-2). Although the hallmark symptoms of COVID-19 are respiratory in nature and related to pulmonary infection, extrapulmonary effects have been reported in COVID-19 patients ^1,2^, including the central nervous system (CNS) ^3^. Notably, over 30% of hospitalized COVID-19 patients manifest neurological and even neuropsychiatric symptoms ^4,5^, and some present some degree of encephalitis at some point ^6^. There are increasing reports of persistent and prolonged effects after acute COVID-19, a long COVID-19 syndrome characterized by persistent symptoms and/or delayed or long-term complications beyond 4 weeks from the onset of symptoms. One of these persistent symptoms are neuropsychiatric sequelae ^3^. One study revealed that more than half of hospitalized patients continue to exhibit neurological symptoms for as long as three months after the acute stage ^7^. Impaired cognition was also confirmed in recovered patients after hospitalization ^8–11^, and neurological impairment is consistent with substantial damage to the nervous system ^12^. Previous studies on Severe Acute Respiratory Syndrome (SARS) patients reported the presence of the SARS coronavirus in the brain tissue and cerebrospinal fluid of subjects who presented neurological symptoms ^13–15^. SARS-CoV-2 RNA was also detected in the cerebrospinal fluid of patients with meningitis ^16–18^. Moreover, alterations in the cerebral cortical region, compatible with viral infection ^19^, and a loss of white matter and axonal injury ^20^ have all been reported in COVID-19 patients.

In line with the potential neurotropic properties of SARS-CoV-2, recent evidence has demonstrated the presence of viral proteins in brain regions of COVID-19 patients as well as in the brains of K18-ACE2 transgenic mice infected with SARS-CoV-2 ^21–23^ and syrian hamsters ^24^. The presence of SARS-CoV-2 in the human brain has been associated with significant astrogliosis, microgliosis and immune cell accumulation ^21^. Further indicating the ability of SARS-CoV-2 to infect cells of the CNS, SARS-CoV-2 has also been shown to infect human brain organoid cells in culture ^22,25–27^. Recently, studies have shown evidence that SARS-CoV-2 can cross the blood–brain barrier (BBB) in mice ^28–30^ and in 2D static and 3D microfluidic *in vitro* models ^31,32^, therefore potentially reducing the integrity of the BBB.

Despite the accumulating evidence, we still lack understanding of the cellular and molecular mechanisms involved in SARS-CoV-2 brain infection and the consequent repercussions on brain structure and functionality. To gain further insight into the neuropathological and neurological consequences of COVID-19 and possible cellular and molecular mechanisms, we performed a broad translational investigation in living patients, *postmortem* brain samples and pre-clinical *in vitro* and *ex vivo* models. Clinical and brain imaging features of COVID-19 patients were found to be associated with neuropathological and biochemical changes caused by SARS-CoV-2 infection in the CNS. We found that astrocytes are the main sites of viral infection within the CNS. SARS-CoV-2-infected astrocytes exhibited marked metabolic changes resulting in a reduction of the metabolites used to fuel neurons and build neurotransmitters. Infected astrocytes were also found to secrete unidentified factors that lead to neuronal death. These events could contribute to the neuropathological alterations and neuropsychiatric symptoms observed in COVID-19 patients.

## RESULTS

### Neuropsychiatric symptoms in convalescent COVID-19 patients correlate with altered cerebral cortical thickness

To explore structural cortical brain damage in COVID-19 patients and dissociate it from the indirect consequences of the severe stage of the disease, we performed a cortical surface-based morphometry analysis (using high-resolution 3T MRI) on 81 subjects diagnosed with mild COVID-19 infection (62 reported anosmia or dysgeusia) who did not require oxygen support. The analysis was performed within an average (SD) interval of 57 (26) days after SARS-CoV-2 detection by RT-qPCR, and the subjects were compared to 145 healthy volunteers scanned before the COVID-19 pandemic (Supplementary Tables 1 and 2). The analysis revealed areas of reduced cortical thickness in the left lingual gyrus; calcarine sulcus, including the cuneus; and olfactory sulcus, including the rectus gyrus (Fig. 1A). In contrast, a thicker cortex was detected in the central sulcus (including the precentral and postcentral gyrus) and superior occipital gyrus (Fig. 1A), which can be associated with vasogenic edema ^33^.

**Figure 1.**
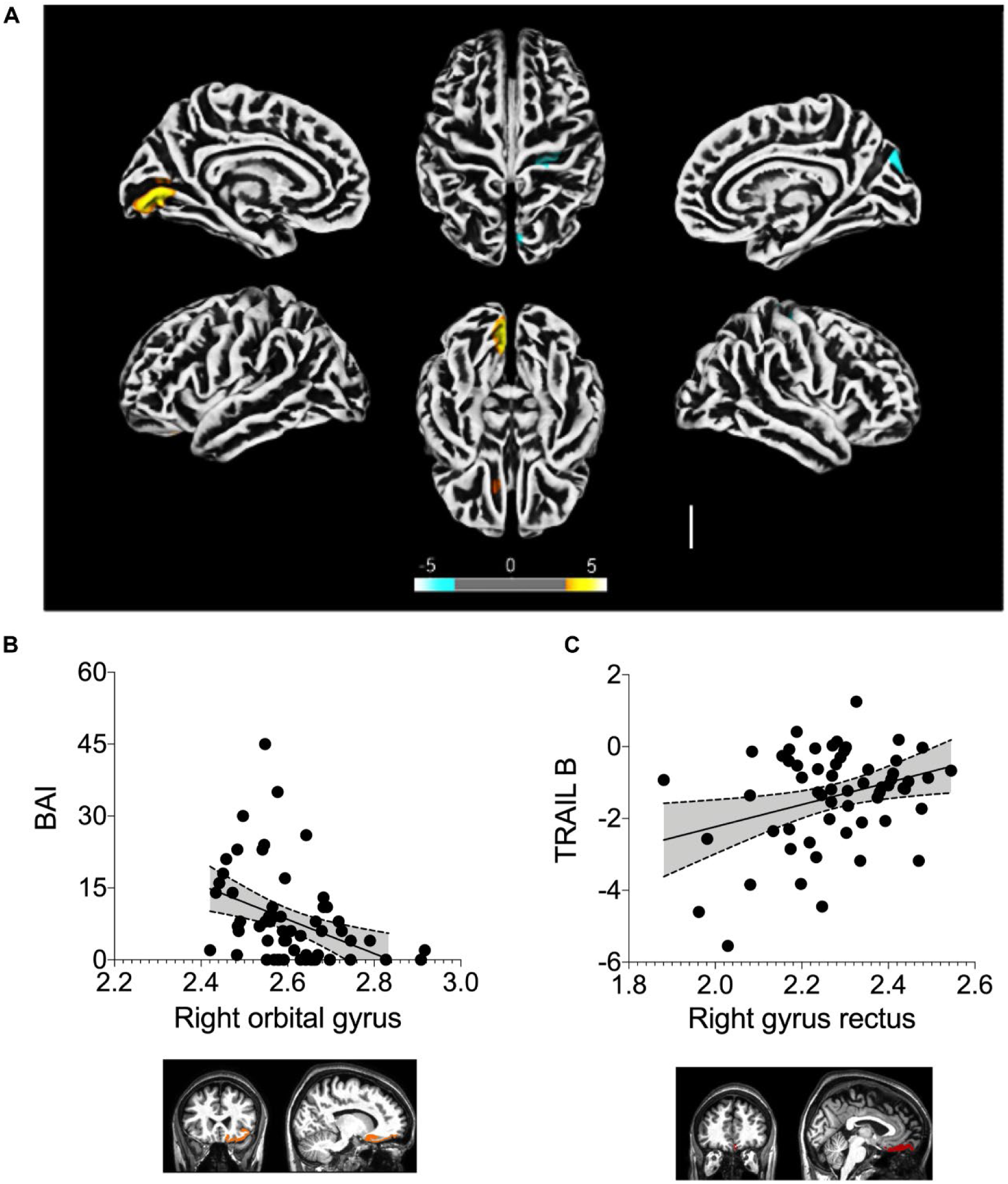
Altered cerebral cortical thickness is associated with neuropsychiatric symptoms in COVID-19 patients. (A) Surface-based morphometry using high-resolution 3T MRI. Yellow represents areas of decreased cortical thickness: left lingual gyrus, calcarine sulcus (and cuneus) and olfactory sulcus (and rectus gyrus). Blue represents areas of increased cortical thickness: central sulcus (precentral and postcentral gyrus) and superior occipital gyrus. Representative image of the analysis of 81 subjects who tested positive for SARS-CoV-2 (who had mild respiratory symptoms and did not require hospitalization or oxygen support) compared to 145 healthy volunteers (without diagnosis of COVID-19). The analysis was performed within an average (SD) interval of 57.23 (25.91) days. (B) Correlation between anxiety scores (BAI) and right orbital gyrus thickness. The data depicts Pearson’s correlation coefficient and region of interest in representative images. (C) Correlation between TRAIL B cognitive performance scores and right gyrus rectus thickness. Data depict Pearson’s correlation coefficient and region of interest in representative images.

A subgroup of 61 participants underwent neuropsychological evaluation, including logical memory (Wechsler Memory Scale) and cognitive function (TRAIL Making Test). Even though these tests were performed a median of 59 days (range between 21-120 days) after diagnosis, we observed fatigue in approximately 70% of individuals and daytime sleepiness in 36%. Nearly 28% of participants performed abnormally on the logical memory cognitive function tests, with approximately 34% and 56% performing abnormally with TRAIL A and B, respectively (Supplementary Fig. 1; Supplementary Table 3). It was interesting that 77% of the COVID-19 patients presented anosmia or dysgeusia in the acute stage, which may be related to the observed changes in cortical thickness ^19^. The high proportion of anosmia and dysgeusia support the idea of the virus entering the nervous system, more specifically the orbitofrontal region (due to the proximity and communication with the nasal cavity). Therefore, alterations in the orbitofrontal cortex are not surprising.

We then interviewed one group of 81 healthy volunteers (HV) (recruited from the same environment as COVID-19 patients) who never presented symptoms or tested positive for COVID-19. Compared to patients (who answered the same question inventories), the HV-group was balanced for age (p=0.99) and sex (p=1). The intensity of symptoms of anxiety (BAI scores) was higher in the COVID-19 group (median of 6, range 0-45) compared to HV (median 1.5, range 0-29), p=0.046. There was also a tendency to have a higher intensity of symptoms of depression (BDI scores) in the COVID-19 group (median 6, range 0-36) compared to HV (median 3, range 0-26), p=0.051. As such, at least part of the long-term neuropsychiatric symptoms manifested by COVID-19 patients can be directly attributed to SARS-CoV-2 infection, even though these patients had only mild respiratory symptoms.

We also observed a negative correlation between BAI scores and the cortical thickness of orbitofrontal regions (adjusted for fatigue scores) (Fig. 1B; Supplementary Table 4), as well as a positive correlation between TRAIL B and cortical thickness of the right gyrus rectus (Fig. 1C; Supplementary Table 5). We additionally identified significant partial correlations between logical memory (immediate recall test, adjusted for anxiety, depression and fatigue) and cortical thickness of regions associated with language (Supplementary Table 6). These results suggest that a thinner cortex in these areas is associated with poor performance on this verbal memory task. Overall, our findings indicate that alterations in the cortical structure are associated with neuropsychiatric symptoms in COVID-19 patients with mild or no respiratory symptoms.

### SARS-CoV-2 infects and replicates in human brain astrocytes of COVID-19 patients

Brain alterations in COVID-19 patients are hypothesized to be a consequence of either inflammatory or hemodynamic changes secondary to peripheral infection, or SARS-CoV-2 invading the CNS and compromising cell viability and brain function. Although exacerbated inflammation and cardiovascular dysfunction have been well-characterized in COVID-19 patients that progress to the severe stages of the disease ^34^, the degree of infection of the CNS by SARS-CoV-2 remains elusive. We performed a minimally invasive autopsy via endonasal trans-ethmoidal access to obtain brain samples from 26 individuals who died of COVID-19. It is worth mentioning that these samples were from brain regions in close proximity to the areas in which we identified altered cortical thickness in the patients of the previous cohort. Initially, we performed an unbiased histopathological analysis of hematoxylin and eosin-stained brain sections to search for features of brain alterations.

We observed alterations consistent with necrosis and inflammation in 5 of the 26 brain tissue samples (Supplementary Table 7 and Table 8). A deeper analysis revealed a strong predominance of senile changes such as corpora amylacea, lipofuscin deposits and parenchymal retraction around the vessels and the meninges. Due to the type of collection performed, alternating white and gray matter was observed. The intraparenchymal inflammatory process was minimal, but present, represented by lymphocytes and perivascular microglia proliferation. In two cases, more intense inflammation was observed, delimiting tiny inflammatory aggregates, associated with endothelial hyperplasia or gemistocytic astrocytes. Cases with nasal epithelium sampled together with brain puncture demonstrated adaptive epithelial changes with cell balonization of the most superficial cells. A few cases showed small, multifocal areas of liquefaction necrosis (Fig. 2A; Supplementary Fig. 2A). It is worth noting that we cannot necessarily discard the cases in which we did not identify tissue alteration; the possibility remains that brain regions different from those at the collection site may still have exhibited brain damage.

**Figure 2.**
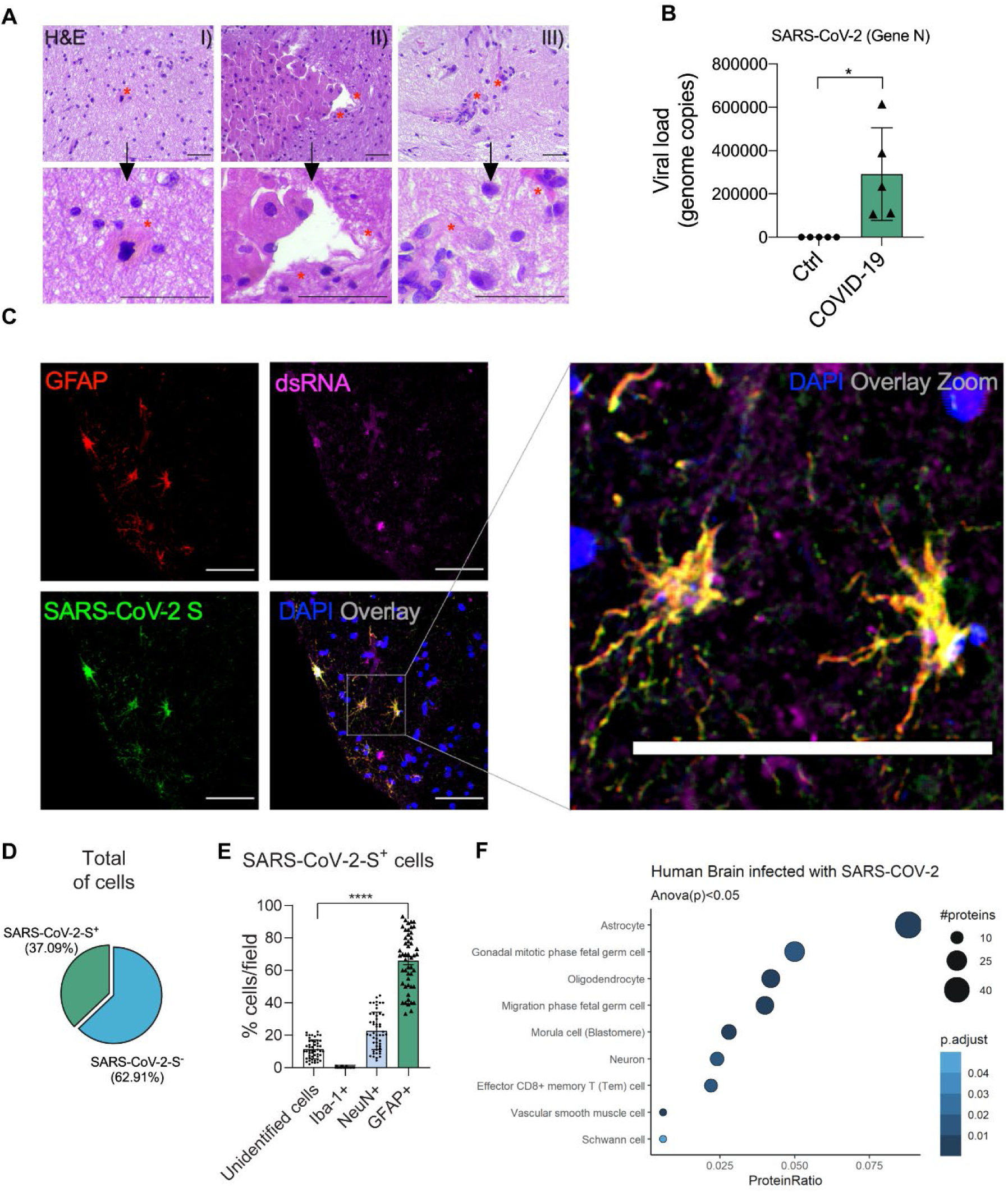
SARS-CoV-2 infects the central nervous system, replicates in astrocytes and causes brain damage. (A) Histopathological H&E images of *postmortem* brain tissue from individuals who died of COVID-19. Five out of 26 individuals showed signs of brain damage as represented in the images by (I) areas of necrosis, cytopathic damage (i.e., enlarged, hyperchromatic, atypical-appearing nuclei), (II) vessels with margination of leukocytes and thrombi and (III) an infiltration of immune cells. The alterations are indicated by red asterisks and a respective zoom. Images were acquired with 400x magnification. Scale bar indicates 50 µm. (B) Viral load in brain tissues from the five COVID-19 patients who manifested histopathological alterations in the brain as compared to samples from SARS-CoV-2-negative controls (n=5 per group). (C) Representative confocal images of the brain tissue of one COVID-19 patient who manifested histopathological alterations. Immunofluorescence targeting glial fibrillary acidic protein (GFAP, red), double-stranded RNA (dsRNA, magenta), SARS-CoV-2-S (green) and nuclei (DAPI, blue). Images were acquired with 630x magnification. Scale bar indicates 50 µm. (D) Percentage of SARS-CoV-2-S positive cells in the tissue of the five COVID-19 patients. (E) Percentage of GFAP-positive vs. unidentified cells, Iba1 and NeuN among infected cells. Ten fields/cases were analyzed. (F) Cell type enrichment analysis using the dataset generated from *postmortem* brain tissue from patients who died of COVID-19. Dot size represents the number of proteins related to the respective cell type and the color represents the p-value adjusted by false discovery rate (FDR). All data are shown as mean ± SEM. P-values were determined by two-tailed unpaired tests with Welch’s correction (B) or ANOVA one-way followed by Tukey’s post hoc test (E).*P < 0.05 compared to the control group; **** P < 0.0001 compared to unidentified cells, Iba1 and NeuN groups.

Next, hypothesizing that the histopathological signs of brain damage would guide us to SARS-CoV-2 brain infection, we evaluated the presence of SARS-CoV-2 in the five brain samples that presented histopathological alterations (Supplementary Table 7; Table 8). Indeed, both SARS-CoV-2 genetic material and spike protein were detected in all five samples (Fig. 2B; Fig. 2C). On average, SARS-CoV-2 spike protein was detected in 37% of the cells in the brain tissue (Fig. 2D), the majority of these SARS-CoV-2 spike-positive cells (65.93%) being astrocytes (GFAP+ cells; Fig. 2E; Supplementary Fig. 2B). As such, approximately 25% of all cells in the brain samples of the five COVID-19 patients that we analyzed were infected astrocytes. We also detected SARS-CoV-2 spike protein in neurons, although to a lesser extent (NeuN+ cells; Fig. 2E; Supplementary Fig. 3A). We did not find signs of infection in microglia (Iba-1+ cells; Fig. 2E; Supplementary Fig. 3B). The specificity of anti-spike antibodies was validated in brain tissue of COVID-19-free cases and in SARS-CoV-2-infected Vero cells (Supplementary Fig. 4A-4C). Additionally, the presence of SARS-CoV-2 spike protein was directly correlated with the presence of double-stranded RNA (dsRNA) in the cells (Fig. 2C; Fig. Supplementary Fig. 3C), indicating the presence of replicative viruses in the brain tissue ^35^.

To investigate the molecular changes at the protein level caused by SARS-CoV-2 infection, we conducted an LC/MS proteomic analysis with a separate set of samples, consisting of 12 *postmortem* brain samples from COVID-19 patients *vs.* 8 SARS-CoV-2-negative controls. We identified 656 differentially expressed proteins, 117 downregulated and 539 upregulated. Pathways associated with neurodegenerative diseases, carbon metabolism and oxidative phosphorylation were enriched in these samples (Supplementary Fig. 5). Notably, astrocytic proteins were the most present among the differentially expressed proteins, consistent with the higher frequency of infected astrocytes observed in COVID-19 *postmortem* brains (Fig. 2F).

To confirm that SARS-CoV-2 infects human brain cells, we analyzed brain slices from human cortices that were exposed to SARS-CoV-2. Both SARS-CoV-2 spike protein and dsRNA were detected in human brain slices 48 hours post-infection (Fig. 3A). The SARS-CoV-2 genome was also detected by RT-PCR in infected brain slices at both 24 and 48 hours post-infection (Fig. 3B). SARS-CoV-2 infection in astrocytes was confirmed by immunostaining the SARS-CoV-2 spike protein and GFAP (Fig.3C). We observed that the majority of the spike-positive cells (5.65% of total cells) were positive for GFAP (58.33% of infected cells; Fig. 3D), similar to the percentage of infected astrocytes observed in the five histologically altered postmortem samples. Taken together, these data indicate that SARS-CoV-2 can preferentially infect astrocytes and replicate within them, in line with human *postmortem* brain findings.

**Figure 3.**
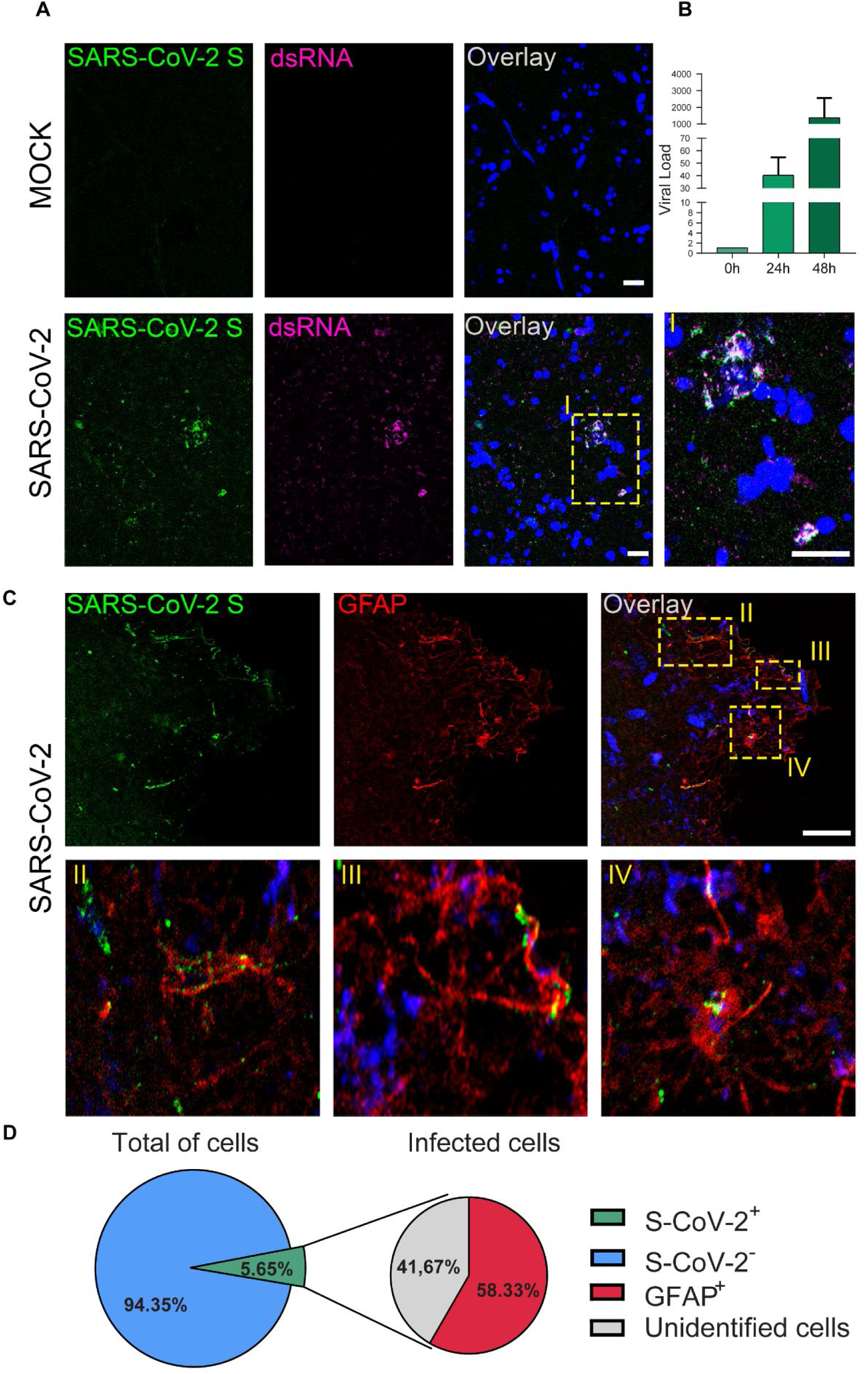
SARS-CoV-2 infects, replicates in and preferentially targets astrocytes in adult human brain slices. (A) Representative images of uninfected control (mock) and SARS-CoV-2-infected human brain slices. Tissue sections were stained for SARS-CoV-2 spike protein (green), double stranded RNA (magenta), and nuclei (DAPI, blue), as indicated. Scale bar indicates 20 µm. Higher magnification is shown in I. (B) Viral load determined by RT-PCR in a human brain slice infected by SARS-CoV-2 at 0, 24 and 48 hours post-infection compared to mock slices (n=2 slices from one patient). (C) Representative cells double-labeled with SARS-CoV-2 spike protein (green) and GFAP (red) are shown at a higher magnification in II, III and IV. (D) Quantification of cells positive for SARS-CoV-2 spike protein and GFAP in human brain slices (n= 2 fields from one patient). Scale bar indicates 20 µm.

To obtain further evidence of the susceptibility of human astrocytes to SARS-CoV-2 infection, neural stem cell-derived human astrocytes (BR-1 cell line) ^36^ were exposed to the virus, and the viral load was determined 24h after infection (Fig. 4A). In agreement with what was observed in the *postmortem* brain samples, we confirmed that SARS-CoV-2 infects human astrocytes (Fig. 4B-4E), as shown by the detection of viral genetic material (Fig. 4B) and spike protein in infected cells (Fig. 4C; Fig. 4D). dsRNA was also observed in SARS-CoV-2-infected astrocytes *in vitro*, but not in mock-infected control cells, suggesting viral replication occurs in these cells (Fig. 4C; Fig. 4E). We also observed that SARS-CoV-2 infection reduced human astrocyte viability by 25% at 48h post-infection, and remained at that reduced viability after 72h (Fig. 4F). To confirm the susceptibility of human astrocytes to SARS-CoV-2 infection, we used a replication-competent pseudotyped vesicular stomatitis virus (VSV) in which the glycoprotein gene (G) of VSV was replaced by the full-length SARS-CoV-2 spike (S) and which contained an eGFP reporter (VSV-eGFP-SARS-CoV-2) ^37^. Our data revealed that the human astrocyte culture was infected by VSV-eGFP-SARS-CoV-2, further supporting the idea that human astrocytes are susceptible to SARS-CoV-2 infection (Fig. 4G; Fig. 4H). Altogether, these results indicate that human astrocytes are permissive cells for SARS-CoV-2 infection and also likely represent a site for viral replication in the CNS.

**Figure 4.**
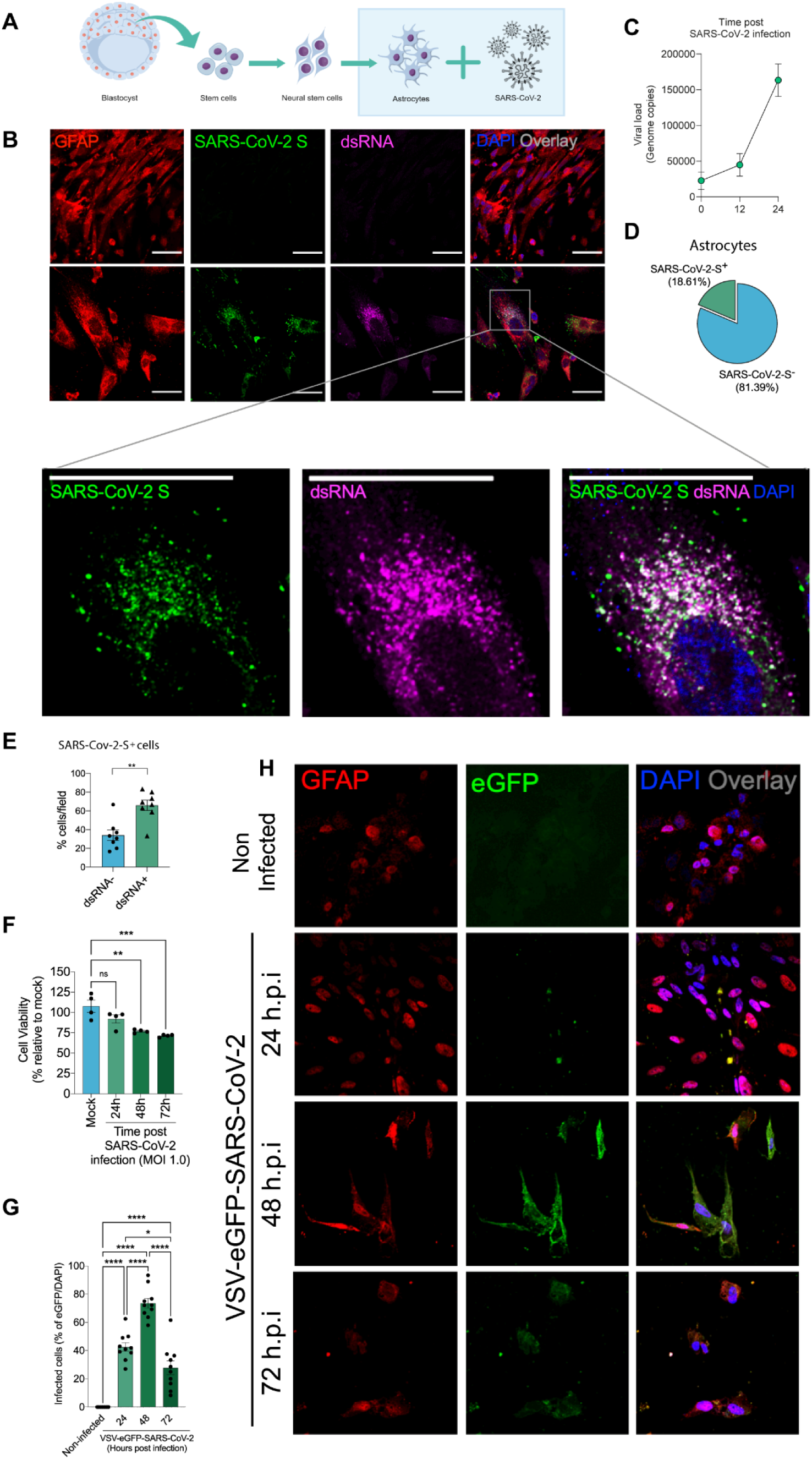
SARS-CoV-2 infects and replicates in astrocytes *in vitro*. (A) Human neural stem cell-derived astrocytes were infected *in vitro* with SARS-CoV-2 (MOI 1.0) for 1h, washed and harvested 24h after infection. (B) Immunostaining for GFAP (red), double-stranded RNA (dsRNA, magenta), SARS-CoV-2-S (green) and nuclei (DAPI, blue). Images were acquired with 630x magnification. Scale bar indicates 50 µm. (C) SARS-CoV-2 viral load detection in astrocyte cell pellets (n = 6 replicates) using RT-PCR. (D) Percentage of infected astrocytes. The data depict SARS-CoV-2-S and DAPI-stained cells (100 fields were analyzed). (E) Frequency of cells containing replicating viruses. (F) Astrocyte viability upon SARS-CoV-2 infection was assessed using a luminescence-based cell viability assay (CellTiter-Glo), determining the number of live cells by quantification of ATP at 24, 48 and 72 hours post-infection. (G) Percentage of infected cells with pseudotyped SARS-CoV-2 (VSV-eGFP-SARS-CoV-2) at 24, 48 and 72 hours post-infection. (H) Staining for DAPI (nuclei, blue), GFAP (astrocytes, red) and eGFP (virus, green) in astrocytes infected with pseudotyped SARS-CoV-2 (VSV-eGFP-SARS-CoV-2) at 24, 48 and 72 hours post-infection. The data represent the percentage of dsRNA-stained cells of SARS-CoV-2-S positive cells (10 fields were analyzed). All data are representative of at least two independent experiments performed in triplicate or quadruplicate and shown as mean ± SEM. P-values were determined by two-tailed unpaired tests with Welch’s correction (E) or one-way ANOVA followed by Tukey’s post hoc test (F and G). **P < 0.01; *** P < 0.001; **** P < 0.0001 compared to the mock group.

### NRP1 is required for infection of astrocytes by SARS-CoV-2

Since astrocytes are susceptible to SARS-CoV-2 infection, we searched for the receptor involved in the viral entry mechanism. To do so, we started by using a publicly available single-cell RNAseq dataset from brain samples of COVID-19 patients ^38^ to analyze the expression of classical SARS-CoV-2 receptors such as ACE2, as well as alternative receptors NRP1 and BSG ^39–41^(Supplementary material). These analyses revealed that ACE2 mRNA was undetected in astrocytes (data not shown) ^21,38^; however, astrocytes did express detectable levels of NRP1 and BSG mRNA (Fig. 5A; Fig. 5B). We also found that the expression of NRP1 and BSG mRNA was increased in astrocytes from COVID-19 patients compared to controls, as well as the percentage of astrocytes expressing these receptors (Fig. 5A; Fig. 5B). Since the binding of SARS-CoV-2 spike to BSG remains controversial ^42^, we decided to explore the possible role of NRP1. Firstly, we performed Western blotting using cultured neural stem cell-derived astrocyte extracts to evaluate if the data from single-cell RNAseq could be confirmed at the protein level in our *in vitro* model. We confirmed that human astrocytes do not express ACE2, whereas they do express NRP1 (Fig. 5C-5E; Supplementary Fig. 6). A549 cells transduced with ACE2 were used as the positive control. To determine the role of NRP1 in the mechanisms behind SARS-CoV-2 infection in astrocytes, we pre-incubated these cells with a neutralizing, anti-NRP1 antibody. Neutralization of NRP1 inhibited the infection of cultured astrocytes by SARS-CoV-2 (Fig. 5D) as well as the infection of cultured astrocytes by VSV-eGFP-SARS-CoV-2 (Fig. 5E; Fig. 5F). These results indicate that SARS-CoV-2 infects human astrocytes via the NRP1 receptor.

**Figure 5.**
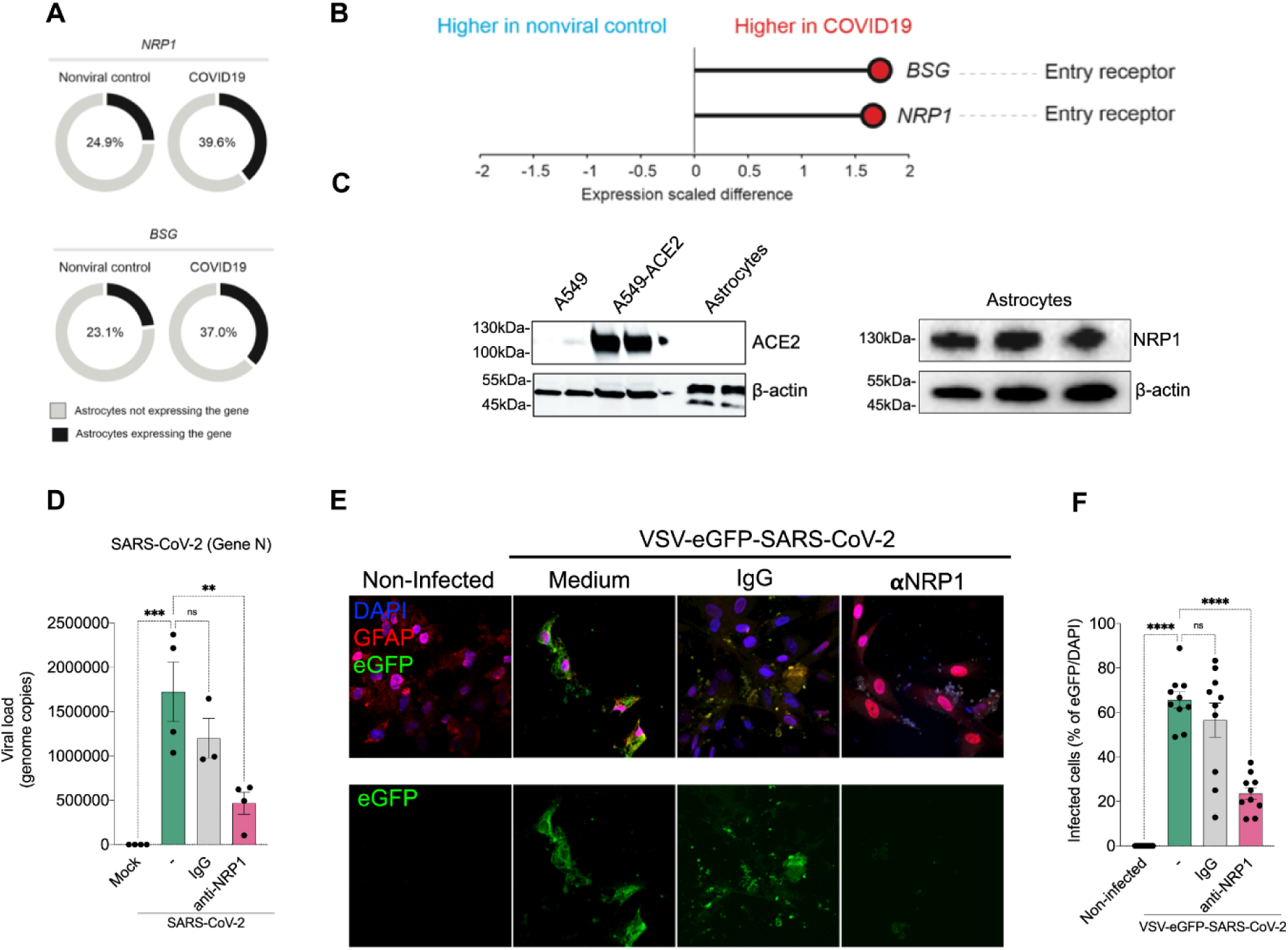
SARS-CoV-2 infects astrocytes via NRP1. (A) Percentage of astrocytes expressing entry receptor genes in COVID-19 patients compared to astrocytes from non-infected controls. (B) BSG and NRP1 are differentially expressed in astrocytes from COVID-19 patients compared to astrocytes from non-infected controls. The x-axis shows the average expression difference (scaled) between COVID-19 patients and non-infected controls. (C) Immunoblot analyses of ACE2 and NRP1 using an extract of non-infected neural stem cell-derived astrocytes. Beta-actin was used as loading control. To control for ACE2 expression, we used A549 cells and A549 cells overexpressing ACE2. (D) Neural stem cell-derived astrocytes were pre-incubated with an NRP1-neutralizing antibody and then harvested 24h post-infection to measure SARS-CoV-2 viral load. (E) Astrocytes were stained for DAPI (nuclei, blue), GFAP (astrocytes, red) and eGFP (virus, green). Cells were pre-incubated with the NRP1-neutralizing antibody and then assessed 48h post-infection with the SARS-CoV-2 pseudotyped virus (VSV-eGFP-SARS-CoV-2). (H) Percentage of infected cells. Images were acquired with 630x magnification. Scale bar indicates 50 µm. All data are representative of at least two independent experiments performed in triplicate and shown as mean ± SEM. P-values were determined by one-way ANOVA followed by Tukey’s post (E and G). **P < 0.01; *** P < 0.001; **** P < 0.0001 compared to the mock group.

### Proteomic and metabolomic changes in SARS-CoV-2-infected human astrocytes

To identify downstream mechanisms that are triggered by SARS-CoV-2 infection and, as such, possibly involved in the changes observed in the brain tissues of COVID-19 patients, we analyzed the proteome of SARS-CoV-2-infected human astrocytes (Fig. 6A). Liquid chromatography-mass spectrometry (LC/MS)-based shotgun proteomics revealed 170 differentially expressed proteins in SARS-CoV-2-infected astrocytes compared to mock-control cells, 68 being upregulated and 102 downregulated (Supplementary Fig. 7). A subset of the differentially expressed proteins showed differences that were sufficient to compose a molecular signature that was able to clearly distinguish the infected astrocytes from mock-controls (Fig. 6A).

**Figure 6.**
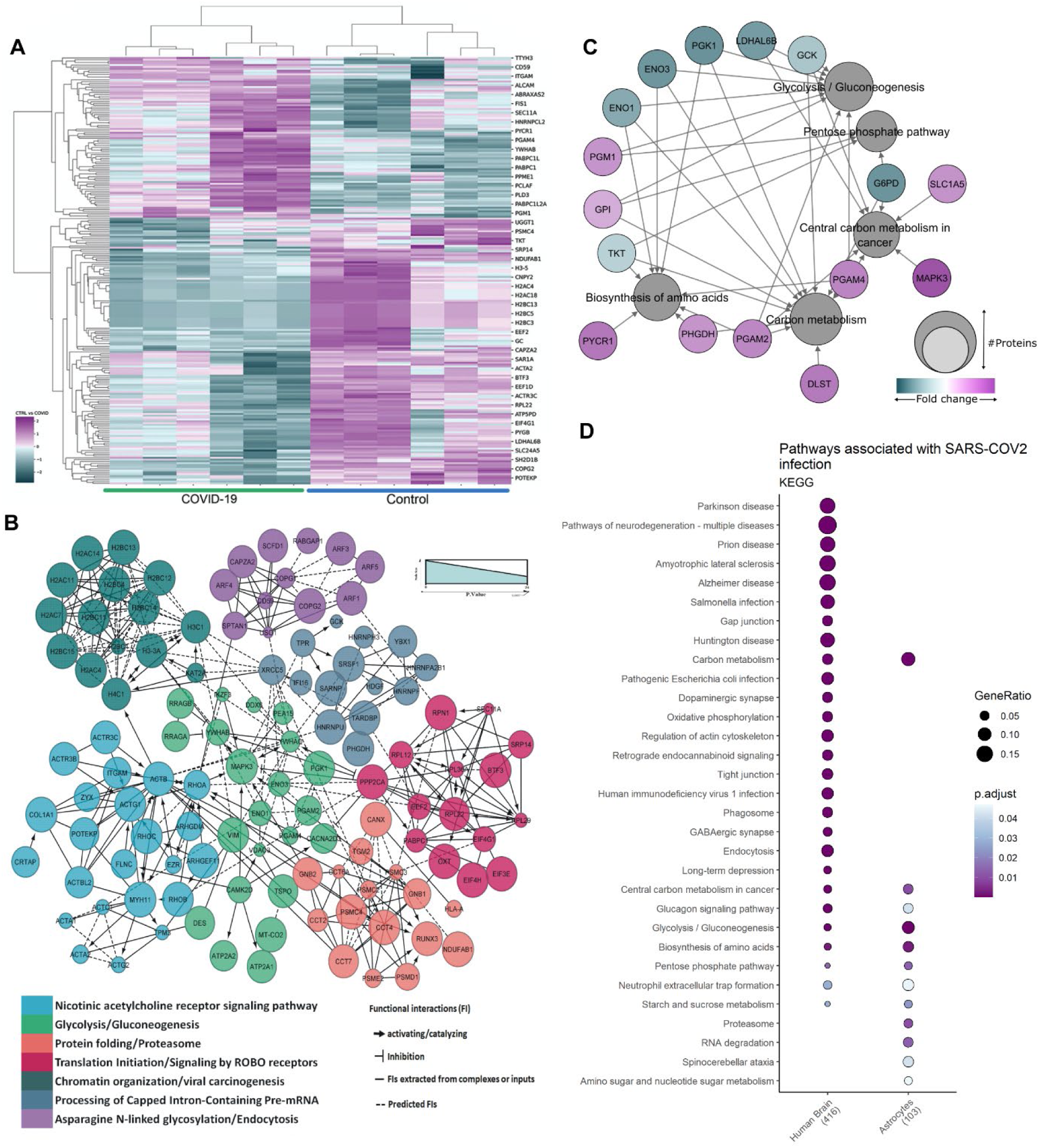
Proteomic changes in SARS-CoV-2-infected human astrocytes and *postmortem* brain tissue from COVID-19 patients. (A) Hierarchical clustering of differentially expressed proteins in human neural stem cell-derived astrocytes that were infected *in vitro* with SARS-CoV-2 (MOI 0.1) for 1h, washed thoroughly and harvested after 24h. Mock infection was used as a control. (B) Reactome functional interaction network of differentially regulated genes in human neural stem cell-derived astrocytes infected with SARS-CoV-2. Seven different colors show 7 protein clusters of enriched pathways and the line types represent protein-protein interactions and downstream activation or inhibition related to gene modulation, showing how some pathways can be affected by SARS-CoV-2 infection (p<0.05 calculated based on binomial test). (C) Network of proteins found differentially regulated in SARS-COV-2-infected astrocytes and their respective pathways, enriched according to the KEGG database. The pathways are represented by gray circles and their size is proportional to the number of proteins differentially regulated; proteins are represented by the smallest circles, colored according to their fold change. (D) Cell type enrichment analysis using the dataset generated from *postmortem* brain tissue from patients who died of COVID-19. Dot size represents the number of proteins related to the respective cell type and the color represents the p value adjusted by false discovery rate (FDR). (E) KEGG enrichment analysis of differentially expressed proteins in SARS-CoV-2-infected astrocytes *vs*. mock as compared to *postmortem* brain tissue from COVID-19 patients *vs.* controls. Dot size represents the number of proteins related to the respective cell type and the color represents the p-value adjusted by false discovery rate (FDR).

Using these same dysregulated proteins, pathway enrichment and interactome analyses predict a wide range of biological processes and regulatory networks to be affected by SARS-CoV-2 infection (Fig. 6B). Pathways involved in carbon/glucose metabolism were among the most enriched (Fig. 6C). More specifically, proteins found differentially expressed in SARS-CoV-2-infected astrocytes and in *postmortem* brain tissue samples belong to glycolysis/gluconeogenesis, carbon metabolism and the pentose phosphate pathway (Fig. 6D). Collectively, these data reinforce that SARS-CoV-2 infects astrocytes in the CNS, possibly affecting energy metabolism pathways and modulating proteins associated with neurodegeneration.

Since there are significant proteomic alterations in metabolic pathways, we sought to investigate if the infection of human astrocytes impacts the levels of key metabolites involved in energy metabolism. LC/MS-based metabolomic analysis of SARS-CoV-2-infected astrocytes showed pronounced changes in metabolic intermediates of glycolysis and anaplerotic reactions, indicating alterations in the pathways of astrocyte metabolism (Fig. 7A). This phenomenon was marked by a decrease in pyruvate and lactate, which are downstream metabolites of the glycolytic pathway, as well as a reduction in glutamine and intermediates of glutamine metabolism such as glutamate, GABA and alpha-ketoglutarate (Fig. 7A). Despite these variations, there were no significant changes in tricarboxylic acid cycle (TCA cycle) intermediates (Supplementary Fig. 8). SARS-CoV-2-infected astrocyte bioenergetics were further characterized by Seahorse Extracellular Flux analysis, showing increased respiration in infected cells (Figure 7B). This was due to an increase in both mitochondrial (maximal respiration) and non-mitochondrial oxygen consumption in SARS-CoV-2-infected astrocytes; the former was linked to higher proton leakage indicating increased uncoupled respiration (Fig. 7B; Fig. 7C). Together, these results demonstrate increased metabolic activity in SARS-CoV-2-infected astrocytes and a reduction of the levels of metabolites produced by these cells to support neuronal metabolism and function.

**Figure 7.**
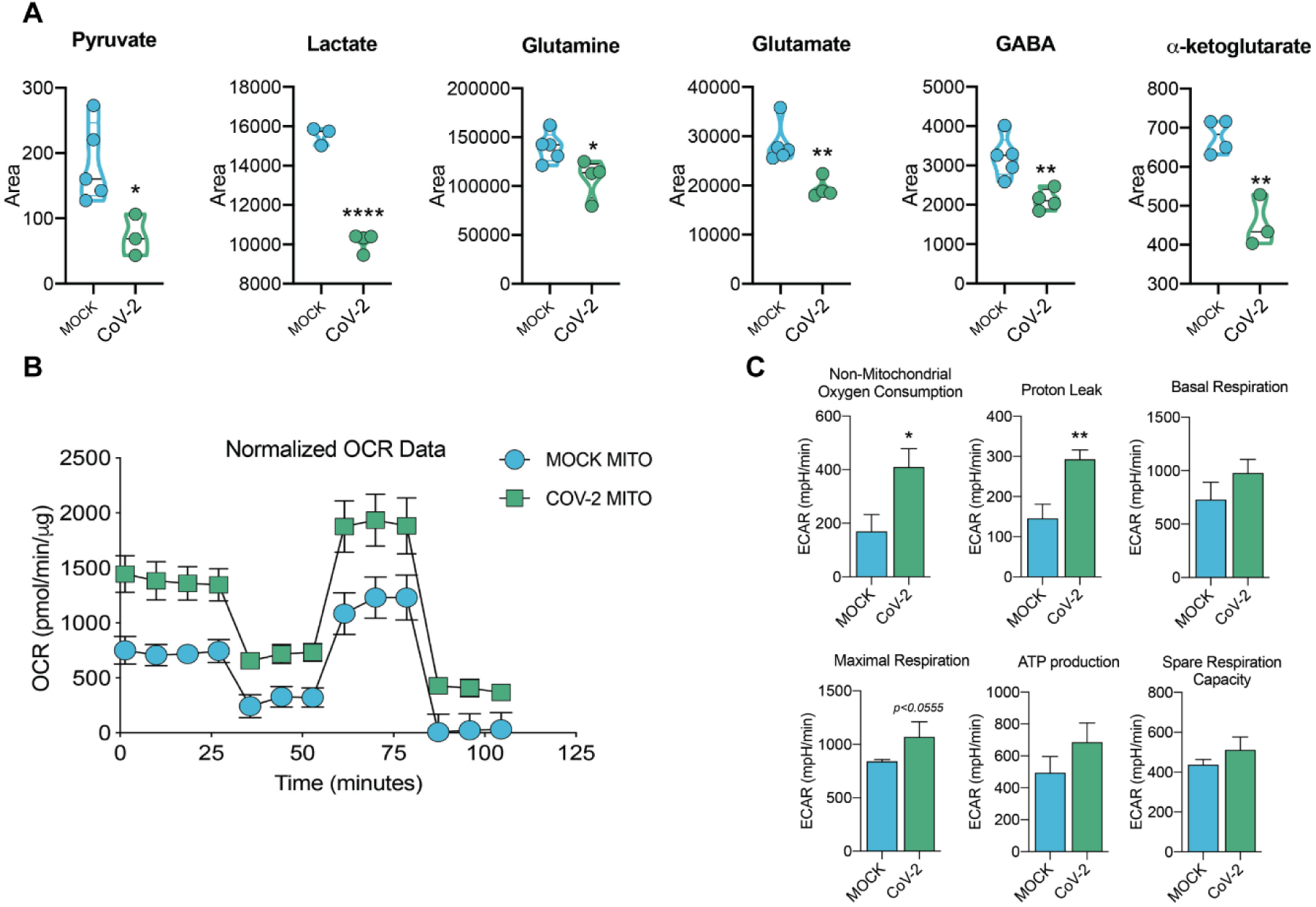
SARS-CoV-2 infection alters astrocyte metabolism. Human neural stem cell-derived astrocytes were infected *in vitro* with SARS-CoV-2 (MOI 0.1) for 1h, washed thoroughly and harvested after 24h. Mock infection was used as a control. (A) High-resolution mass spectrometry-based quantification of pyruvate, lactate, glutamine, glutamate, GABA and ⍺-ketoglutarate in SARS-CoV-2 infected astrocytes vs. mock. The integration area of each peak was used to calculate the violin plot graph and an unpaired t-test with Welch’s correction was used for statistical comparison. (B) Oxygen consumption rate (OCR) of SARS-CoV-2 infected astrocytes *vs.* mock. SeaHorse Flux Analysis using the MitoStress test where basal respiration was measured followed by determination of respiration in response to oligomycin, FCCP and rotenone/antimycin. Data are representative of at least two independent experiments performed in triplicate (metabolomics) or quintuplicate (SeaHorse Flux analysis), and shown as mean ± SEM. P-values were determined by two-tailed unpaired t-test with Welch’s correction. *P < 0.05; **P < 0.01; **** P < 0.0001 compared to mock.

### Conditioned medium from SARS-CoV-2-infected astrocytes reduces neuronal viability

Astrocytes are essential to the control of brain homeostasis, not only because they are the main energy reservoirs of the brain ^43^ but also due to their important role in protective responses to cell damage triggered by infection or sterile inflammation ^44,45^. There is evidence that astrocytes secrete yet-undetermined neurotoxic factors ^44–46^ and are also involved in the uptake, synthesis and distribution of brain metabolites ^47,48^. Thus, we investigated if neuronal viability could be affected by exposure to media conditioned by SARS-CoV-2-infected astrocytes. To test this hypothesis, we cultured either NSC-derived neurons or differentiated SH-SY5Y neurons for 24h in a control or conditioned medium, in which SARS-CoV-2-infected astrocytes were grown (Fig. 8A). The conditioned medium increased the rate of apoptosis by 45.5% and 22.7% in NSC-derived neurons and SH-SY5Y neurons, respectively (Fig. 8B-8E, Supplementary Fig. 9A-9B). This is despite the fact that neither NSC-derived neurons nor SH-SY5Y neurons presented a reduction in cell viability after 24h (Fig. 8D-8E), 48h or 72h (Supplementary Fig. 9C-9D) of being directly exposed to SARS-CoV-2. Ruling out neuronal death by infectious particles in the conditioned medium, SARS-CoV-2 genetic material was not detected in SH-SY5Y neurons or NSC-derived neurons after these cells were incubated with SARS-CoV-2-infected astrocyte-conditioned medium (Supplementary Fig. 9E-F). These results therefore suggest that SARS-CoV-2-infected astrocytes release soluble factors which reduce neuronal viability.

**Figure 8.**
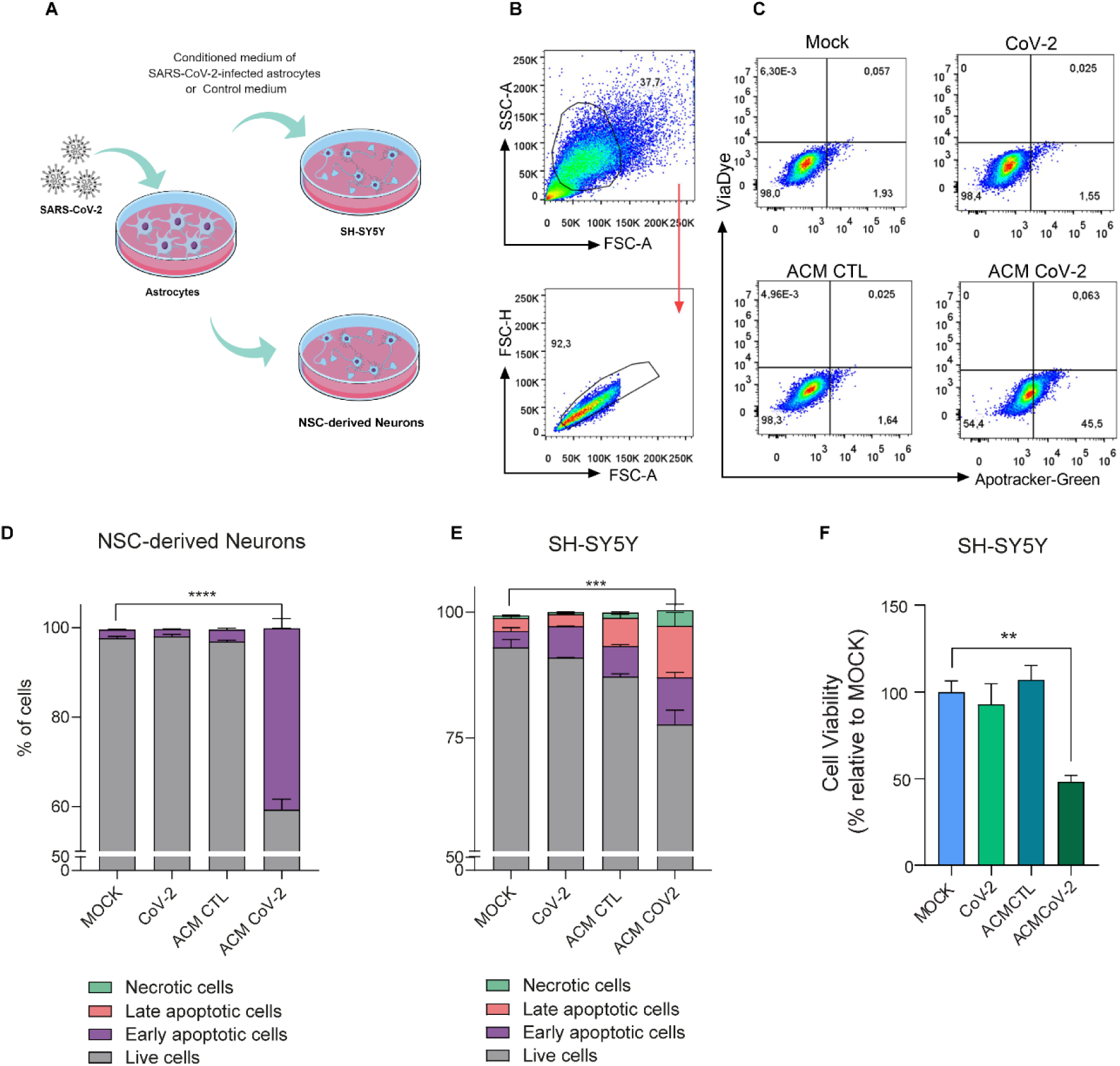
Medium conditioned by SARS-CoV-2-infected astrocytes reduces neuronal viability. (A) Human NSC-derived neurons and SH-SY5Y neuronal cells were cultured for 24h in the presence of medium conditioned by uninfected astrocytes (mock infection; ACM CTL) or SARS-CoV-2-infected astrocytes (ACM CoV-2). (B) Cellular viability as measured by apotracker/fixable viability stain (FVS) and analyzed by flow cytometry. Representative gating strategies are shown. (C) Representative dot-plots of neuronal viability. (D) Percentage of living or nonliving NSC-derived neurons and (E) SH-SY5Y cells. Cells were classified as living (gray bars; double-negative), in early apoptosis (purple bars; apotracker+/FVS-) in late apoptosis (pink bars; double-positive) or necrotic (green bars apotracker-/FVS+). (F) Differentiated SH-SY5Y cells were cultured for 24h in the presence of medium conditioned by SARS-CoV-2-infected astrocytes (ACM CoV-2) or uninfected astrocytes (ACM CTL). The SH-SY5Y cells viability were assessed using an ATP-quantifying, luminescence-based cell viability assay (CellTiter-Glo) at 24h post infection. P-values were determined by one-way ANOVA followed by Tukey’s post hoc test. **P < 0.01; ***P < 0.001; ****P < 0.0001 compared to the mock group.

## DISCUSSION

Our study demonstrates structural and functional alterations in the brain tissue of COVID-19 patients which correlate with neuropsychiatric and neurological dysfunctions. This study and other reports showing alterations in brain structure and the manifestation of neurological symptoms in COVID-19 patients ^49–51^ raise a debate on whether these clinical features are a consequence of peripheral changes or the potential ability of the virus to invade the CNS. Our findings support the latter, at least in part, as we detect SARS-CoV-2 in brain tissue collected from patients who died of COVID-19. SARS-CoV-2 has also been demonstrated to be able to infect brain cells using *in vitro* models such as stem cell-derived neural cells and cerebral organoids ^22,25–27,52^. Viral particles have also been found in the brain ^21^, localized to the microvasculature and neurons ^22^, as well as in the choroid plexus ^53^ and meninges ^54^; however, the magnitude of this infection and its distribution within the brain tissue had not been determined. Here we show that astrocytes are the main locus of infection - and possibly replication - of SARS-CoV-2 in the brains of COVID-19 patients, as evidenced by the detection of the viral genome, the SARS-CoV-2 spike protein and dsRNA in *postmortem* brain tissue, *ex vivo* brain slices and *in vitro* astrocytes. These findings are in line with published evidence showing that astrocytes from primary human cortical tissue and stem cell-derived cortical organoids are susceptible to SARS-CoV-2 infection ^22,52,55^.

Recently, Meinhardt *et al.* 2021 described that SARS-CoV-2 could access the CNS through the neural-mucosal interface in olfactory mucosa, entering the primary respiratory and cardiovascular control centers in the medulla oblongata. Other proposed routes of SARS-CoV-2 neuro-invasion are through the infection of brain endothelial cells. In addition to the inflammatory response produced by SARS-CoV-2 infection, endothelial cell infection could cause dysfunctions in BBB integrity and facilitate access of the virus to the brain ^28–30,32,56,57^. Despite the advances that have been already made, there is still much left to be learned about the routes that SARS-CoV-2 can take to invade the brain and how the virus navigates across different brain regions.

While ACE2 is the best-characterized cellular receptor for the entry of SARS-CoV-2 into cells through the interaction with the viral spike protein, other receptors have also been identified as mediators of infection ^58^. According to our data and others ^21^, astrocytes do not express ACE2, although we found higher expression of NRP1. This receptor is abundantly expressed in the CNS ^39,40,59^, particularly in astrocytes (Fig. 5C); blocking access to it using neutralizing antibodies reduced SARS-CoV-2 infection in these cells. These results indicate that SARS-CoV-2 infects *in vitro* astrocytes via the NRP1 receptor, though this has yet to be confirmed *in vivo*.

To understand the consequences of SARS-CoV-2 infection in NSC-derived astrocytes, we searched for changes in the proteome in a non-hypothesis-driven fashion. SARS-CoV-2 infection resulted in substantial proteomic changes in several biological processes, including those associated with energy metabolism, in line with previous reports on other cell types infected with SARS-CoV-2 ^60–62^. Noteworthily, differentially expressed proteins in COVID-19 *postmortem* brains were enriched for astrocytic proteins, strengthening our own findings that these are the most affected cells by SARS-CoV-2 infection in the human brain. Differentially expressed proteins in COVID-19 *postmortem* brains were also enriched for markers of oligodendrocytes, neurons and Schwann cells, although less significantly than astrocytes. Our proteomic data also evidenced changes in components of carbon metabolism pathways, particularly glucose metabolism, in both *in vitro* infected astrocytes and *postmortem* brain tissue from COVID-19 patients.

Since astrocyte metabolism is key to support neuronal function, changes in astrocyte metabolism could indirectly impact neurons. One way astrocytes support neurons metabolically is by exporting lactate ^63^, and one of the most critical alterations seen here caused by SARS-CoV-2 infection in astrocytes was a decrease in pyruvate and lactate levels. Moreover, intermediates of glutamine metabolism such as glutamate and GABA were decreased in SARS-CoV-2-infected astrocytes. This said, there were no significant changes in any core intermediates of the TCA cycle. Together with the increased oxygen consumption rate of SARS-CoV-2-infected astrocytes, these results suggest that glycolysis and glutaminolysis are being used to fuel carbons into the TCA cycle to sustain the increased oxidative metabolism of infected astrocytes. Astrocyte-derived lactate and glutamine are required for neuronal metabolism ^48,64^ and the synthesis of neurotransmitters such as glutamate and GABA ^65^, respectively. Astrocytes also play a vital role in neurotransmitter recycling, a crucial process for the maintenance of synaptic transmission and neuronal excitability. This is especially important at glutamatergic synapses, since proper glutamate uptake by astroglia prevents the occurrence of excitotoxicity ^66^. Upon this uptake, glutamine synthetase converts glutamate to glutamine, which can then be transferred back to neurons, thus closing the glutamate-glutamine cycle ^67^. This is also true for GABAergic synapses, where the neurotransmitter GABA is taken up by astrocytes and metabolized first to glutamate and then to glutamine ^68^. Moreover, astrocytes are responsible for maintaining glutamate levels in the brain ^69,70^. Recently, de Oliveira *et al.* showed that glutamine levels were reduced in mixed glial cells infected with SARS-CoV-2 and that the inhibition of glutaminolysis decreased viral replication and pro-inflammatory response ^24^. Hence, given the importance of the coupling between astrocytes and neurons, alterations in astrocytic glucose and glutamine metabolism are expected to compromise neuronal function, affecting neuronal metabolism and synaptic function and plasticity ^71^.

In addition to the metabolic changes observed in SARS-CoV-2-infected astrocytes that may impact neuronal dysfunction, we also found that SARS-CoV-2 infection elicits a secretory phenotype in astrocytes that results in increased neuronal death. Our data point to the release of one or more still-unidentified neurotoxic factors by SARS-CoV-2-infected astrocytes. A similar phenomenon has been observed when astrocytes are activated by inflammatory factors ^44,72,73^. Neuronal death may explain, at least partially, the alterations in cortical thickness we observed in COVID-19 patients. A recent study with 60 recovered patients and 39 healthy controls also identified gray matter abnormalities 97 days after the onset of the disease, with increased volume in some areas of the brain ^7^. While that study analyzed hospitalized patients, we evaluated individuals that did not require hospitalization (i.e., had mild respiratory symptoms); nonetheless, we observed notable alterations in cortical thickness. Importantly, some of these alterations correlated with the severity of symptoms of anxiety and impaired cognition, which is consistent with previous literature ^51,74,75^.

It is also worth noting that these altered regions are anatomically close to the brain regions from which we obtained *postmortem* samples (through endonasal trans-ethmoidal access) and identified histopathological alterations associated with SARS-CoV-2 infection. Our findings are consistent with a model in which SARS-CoV-2 is able to reach the CNS of COVID-19 patients, infect astrocytes and secondarily impair neuronal function and viability. These changes may contribute to the alterations of brain structure observed here and elsewhere, thereby resulting in the neurological and neuropsychiatric symptoms manifested by some COVID-19 patients. Our study comes as a cautionary note that interventions directed to treat COVID-19 should also envision ways to prevent SARS-CoV-2 invasion of the CNS and/or replication in astrocytes.

## Materials and Methods

### Brain imaging and neuropsychological evaluation

#### Participants

Eighty-one patients (60 women, median 37 years of age) previously infected with SARS-CoV-2 were enrolled prospectively for this study after signing an informed consent form approved by the local ethics committee. These individuals were diagnosed during the first semester of 2020, presented mild symptoms during the acute phase and did not require hospitalization or oxygen therapy. There was a median interval of 54 days (range 16-120 days) between their RT-PCR exam and the day of MRI scanning and interview. For cortical thickness analysis, we included one hundred and forty-five controls (103 women, median 38 years of age) ^76^ from our neuroimaging databank, given the difficulties and risk of recruiting healthy volunteers during the pandemic. The outpatients and healthy controls were balanced for age (p=0.45) and sex (p=0.65). The neuropsychological evaluations and neuroimaging analyses were approved by the Research Ethics Committee of the University of Campinas (CAAE: 31556920.0.0000.5404) and all subjects signed a consent form to participate.

#### Neuropsychological evaluation

We performed neuropsychological evaluations of sixty-one of these patients. They were tested for symptoms of anxiety using the Beck Anxiety Inventory (BAI) and symptoms of depression using the Beck Depression Inventory (BDI) ^77^. Symptoms of anxiety were considered for those with a BAI higher than 10 points, and depression symptoms defined for those with a minimum of 14 points on the BDI. Anxiety symptoms were classified as mild (BAI 11-19), moderate (20-30) or severe (31-63) and depression symptoms were classified as mild (BDI 14-19), moderate (20-35) or severe (36-63). We evaluated verbal memory (immediate and delayed episodic memory) using the Logical Memory subtest from the Wechsler Memory Scale (WMS-R) ^78^, in which the examiner verbally presents two stories, each including 25 pertinent pieces of information. Subjects are required to recall details of each story immediately after its presentation and again after 20 minutes. To evaluate other cognitive functions, we applied the TRAIL Making Test (TMT) ^79^, which is subdivided into two steps. *Step A* assesses processing speed and visual search in a task that requires connecting 25 randomly arranged numbers in ascending order. *Step B* evaluates alternating attention and cognitive flexibility in a task associated with shifting rules in an ascending sequence of 25 numbers. A training stage is applied to both steps. We calculated z-scores for the results of these tests based on Brazilian normative data ^78,80^. For each test, the function was categorized as preserved (z-score > -0.99, including average, high average, above average and exceptionally high scores); low average score (z-score between -1 and -1.5); below-average score (z-score between -1.51 and -2); and exceptionally low score (z-score < -2) ^81,82^. The Chalder Fatigue Questionnaire (CFQ-11) ^83^ was used to evaluate fatigue in these subjects in which they were instructed to answer 11 questions (measured on a Likert scale 0-3), which yields a global score out of 33.

#### Neuroimaging analysis

We obtained the structural, 3D, T1-weighted images from a 3T Achieva-Philips MRI scanner (voxel size: 1×1×1 mm, TE = 3.2 ms, TR = 7 ms, matrix = 240×240×180, flip angle = 8 and FOV = 240×240 mm^2^) ^76,77^. We performed imaging analyses with the CAT12 toolbox (http://www.neuro.uni-jena.de/cat/, version r1711) within SPM12 (http://www.fil.ion.ucl.ac.uk/spm/, version 7487) using MATLAB 2017b to extract Cortical Thickness (CT) maps, according to the default parameters. The T1 images were first spatially registered and segmented into gray matter, white matter and cerebrospinal fluid before calculating cortical thickness using the projection method described by Dahnke *et al.* ^84^. For voxelwise analysis of extracted maps, we used CAT12/SPM12 tools for an independent t-test (comparing COVID-19 patients and healthy controls), including age and sex as covariates. The results displayed were corrected for multiple comparisons using false discovery rate (FDR) ^85^ correction (p<0.05). For anatomical identification, we used the Destrieux Atlas (2009) ^86^. Cortical parcellation was performed with standard CAT12 tools to extract the cortical thickness of regions of interest for correlations with neuropsychological scores. We used SPSS 22 for statistical analysis of clinical and neuropsychological variables. The FDR procedure was applied to adjust p-values for multiple comparisons (when necessary) with R software ^87^.

### Postmortem brain samples from COVID-19

Twenty-six individuals who died from complications related to COVID-19 were autopsied with an ultrasound-guided, minimally invasive approach using endonasal trans-ethmoidal access, performed within 1 hour of the patient’s death. Brain tissue samples were collected and fixed using a 10% neutral buffered formalin solution. After fixation, the tissue was embedded in a paraffin block and sectioned into slices with a thickness of 3μm. The sections were stained by H&E and immunofluorescence was observed. For proteomic analysis, twelve COVID-19 patients were autopsied using the same approach and the samples were frozen at -80°C. Brain tissue samples were macerated in a lysis buffer (100mM Tris-HCl, pH 8.0, 150mM NaCl, 1mM EDTA, 0.5% Triton X-100) prior to trypsin digestion. The clinical data of all COVID-19 patients and control patients are described in Supplementary Table 7, 8 and 9) The autopsy studies were approved by the National Commission for Research Ethics (CAAE: 32475220.5.0000.5440 and CAAE: 38071420.0.1001.5404).

### Generation of human astrocytes (hES-derived)

Differentiation of glial progenitor cells was performed from neural stem cells (NSC) derived from pluripotent human embryonic stem cells (hES, BR-1 cell line) ^36^, according to the method published by Trindade, 2020 ^88^. NSCs were cultured in plates coated with Geltrex Matrix (Thermo Fisher Scientific, MA, USA) using 1:1 Neurobasal™/Advanced DMEM/F12 medium and 2% neural induction supplement. Upon reaching 50% confluence, the medium was changed to DMEM/F12 (Dulbecco’s Modified Eagle Medium/F12), 1% N2 supplement, 1% fetal bovine serum (FBS) and 1% penicillin-streptomycin, and the plates were maintained at 37°C in humidified air with 5% CO_2_ for 21 days. At this stage, cells were considered glial progenitor cells (GPCs). Subsequently, GPCs were plated at low density (30-40% confluence) on Geltrex-coated plates and treated with DMEM/F12 medium, 1% GlutaMAX Supplement, 10% FBS and 1% penicillin-streptomycin. The differentiation medium was replaced every 2-3 days. After 4 weeks of differentiation, the cells were considered mature astrocytes. These cells were plated on Geltrex-coated coverslips at a density of 4×10^4^ cells per well for immunostaining assays (24-well plates) and 2.5×10^5^ cells per well for viral load and proteomic and metabolomic analysis (6-well plates). All products used for cell culture are from Thermo Fisher Scientific, MA, USA. The characterization of the BR-1 lineage as astrocyte cells has been previously described elsewhere ^36,88–90^. We generated eight batches of human astrocytes from BR-1-derived NSCs. The NSCs were of different passages and were used as biological replicates in independent experiments. Our internal control showed that about 97% of the neural stem cell-derived astrocytes in culture expressed GFAP, 80.4% expressed vimentin and 12.9% expressed SOX-2 (markers of progenitor cells). The neural stem cell-derived astrocyte culture expressed more astrocytic markers than progenitor cell markers, showing an excellent effectiveness of human astrocyte generation (Supplementary Fig. 10).

### Virus strain

The HIAE-02-SARS-CoV-2/SP02/human/2020/BRA virus strain (GenBank accession number MT126808.1) was used for all *in vitro* experiments. The virus was isolated from the first confirmed case of COVID-19 in Brazil and was kindly donated by Prof. Dr. Edison Durigon (ICB-USP). The replication-competent pseudotyped vesicular stomatitis virus with the full-length SARS-CoV-2 spike protein replacing the virus glycoprotein gene, and containing an eGFP reporter (VSV-eGFP-SARS-CoV-2) was engineered and donated by Prof. Dr. Sean P.J. Whelan (Department of Medicine, Washington University School of Medicine, St. Louis, MO, USA) for SARS-CoV-2 entry experiments ^37^. Viral stock was propagated in Vero CCL-81 cells (ATCC), cultivated in DMEM supplemented with 10% heat-inactivated FBS and 1% penicillin and streptomycin (Gibco, Walthmam, MA, USA) and incubated at 37°C with 5% atmospheric CO_2_. Viral titer was determined by the plaque-forming assay using Vero cells.

### In vitro *infection*

Astrocytes were infected with SARS-CoV-2 for 1h using an MOI of 0.1 (proteomics; metabolomics; gene expression, viral load and bioenergetics assays; and flow cytometry analysis) or 1.0 (proteomics and immunostaining) under gentle agitation at room temperature. After viral adsorption, cells were washed twice with phosphate-buffered saline (PBS) and incubated with DMEM/F12 supplemented with 10% FBS, 1% GlutaMAX and 1% penicillin and streptomycin for 24h at standard culture conditions (37°C and 5% atmospheric CO_2_). SARS-CoV-2 entry experiments were performed using SARS-CoV-2 (MOI 1.0) and VSV-eGFP-SARS-CoV-2 pseudotyped particles (MOI 1.0) in the presence of an NRP1-neutralizing antibody (BD Bioscience, Cat. 743129, Clone U21-1283). We used anti-IgG2b as a control antibody (Biolegend, Cat. 406703, Clone RMG2b-1). Full details of the metabolomic, proteomic, gene expression, viral load, flow cytometry and bioenergetics assays, as well as immunostaining details are provided in the Supplementary Material.

## Data Availability

The mass spectrometry proteomic data have been deposited to the ProteomeXchange Consortium via the PRIDE partner repository with the dataset identifier PXD023781 and 10.6019/PXD023781.

## Supplementary Figure

**Supplementary Figure 1.**
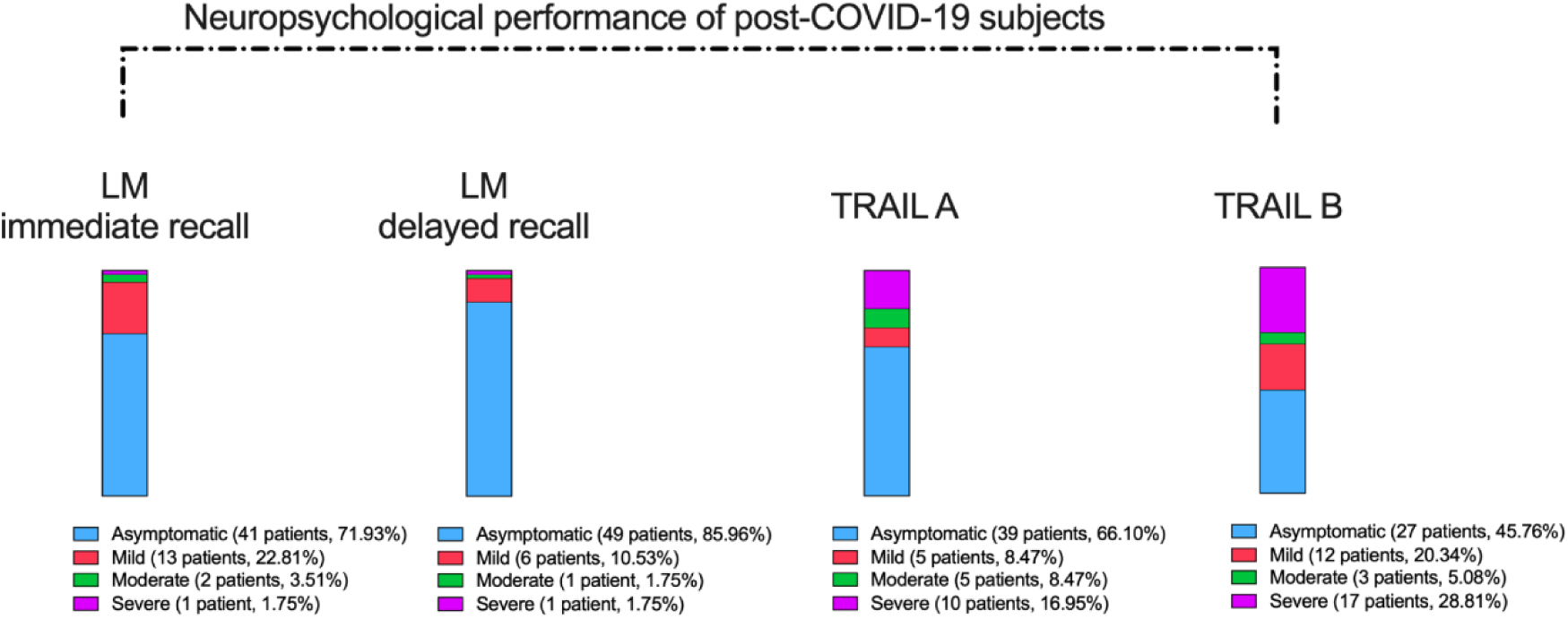
Frequency of neuropsychological impairment in individuals after mild infection. The subgroup of individuals who underwent neuropsychological evaluation presented a median age of 37.8 years (range 21-63 years), 16 years of education (range 6-24 years) and 59 days after COVID-19 diagnosis (range 21-120 days). Severity of impairments in logical memory (LM; Wechsler Memory Scale) and cognitive function (TRAIL Making Test A and B).

**Supplementary Figure 2.**
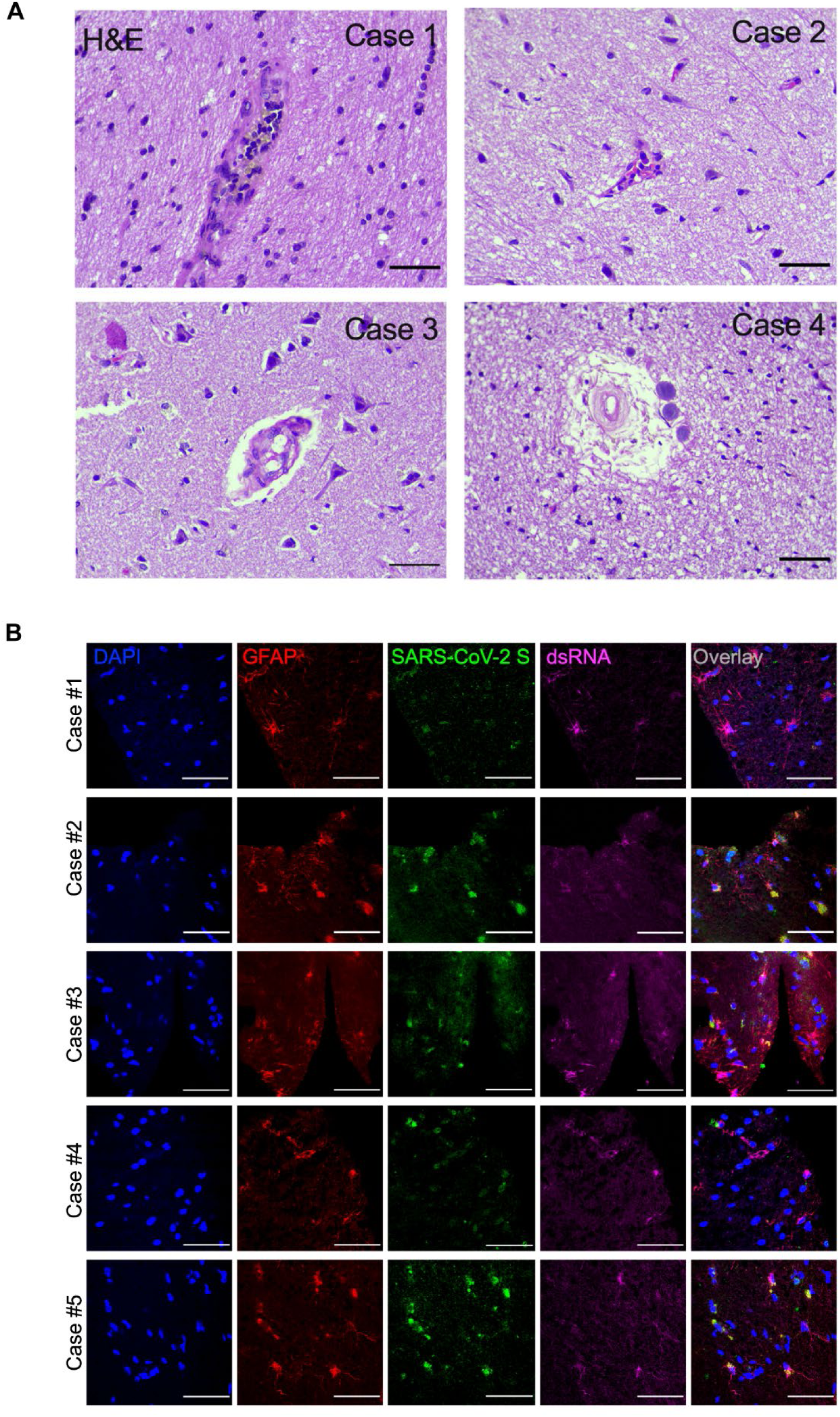
SARS-CoV-2 infects the central nervous system and replicates in astrocytes. (A) Histopathological alterations revealed by H&E images of *postmortem* brain tissue from individuals who died of COVID-19. Samples from 26 individuals were analyzed; 5 showed alterations. Case 1: intraparenchymal cerebral vessel with margination of inflammatory cells through endothelium; Case 2: focal infiltration of inflammatory cells – diapedesis; Case 3: intraparenchymal vascular damage; Case 4: perivascular edema and senile corpora amylacea. Case 5 is also shown in Figure 2. (B) Representative confocal images of the brain tissue of the 5 COVID-19 patients who manifested histopathological alterations. Staining of glial fibrillary acidic protein (GFAP, red), double-stranded RNA (dsRNA, magenta), SARS-CoV-2-S (green) and nuclei (DAPI, blue). Images were acquired with 630x magnification. Scale bar indicates 50µm.

**Supplementary Figure 3.**
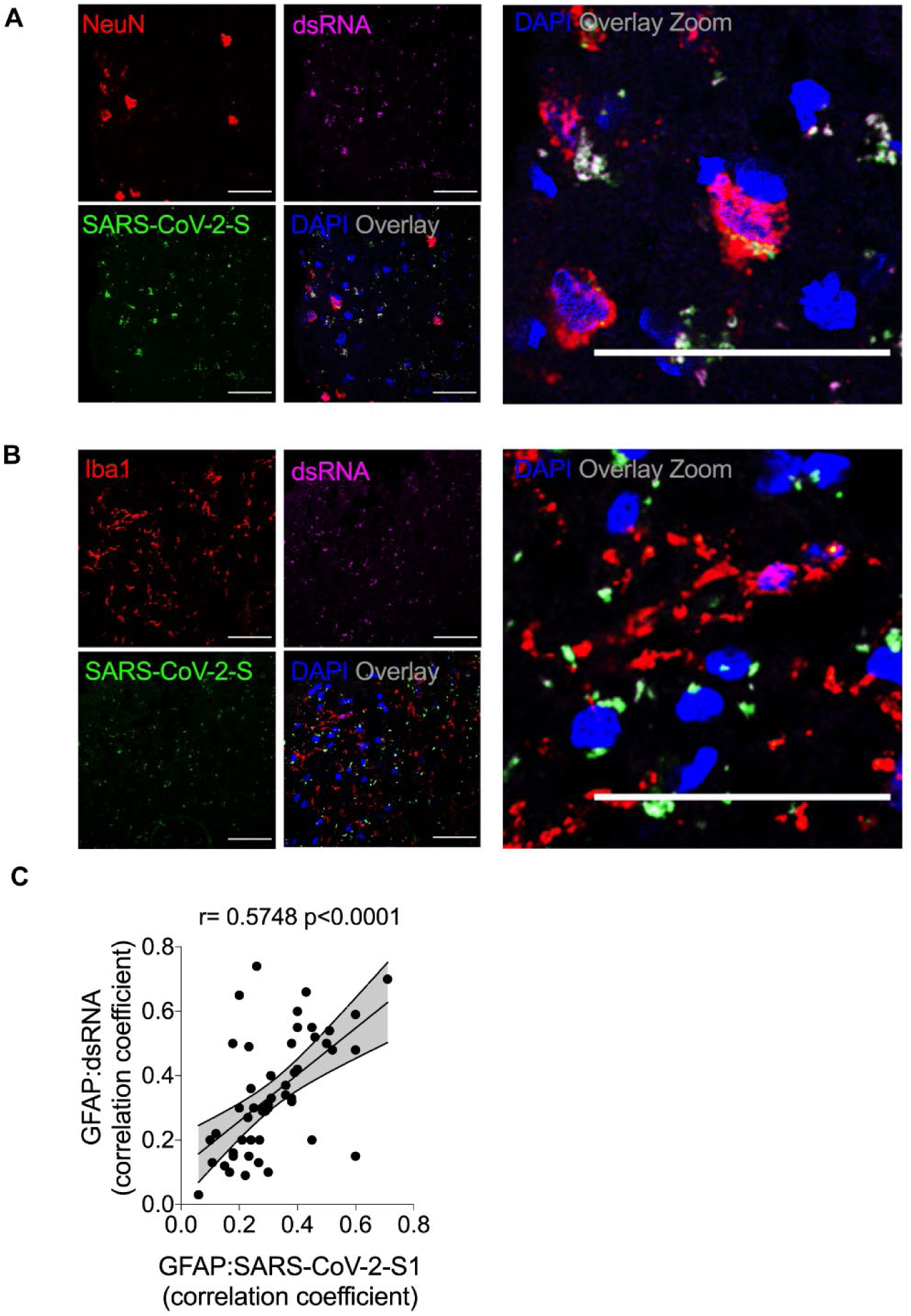
SARS-CoV-2 infects neurons, but not microglia, in the brains of COVID-19 patients. (A-B) Representative immunostaining and confocal analysis from brain slices from autopsies of COVID-19 patients (n=5). The image depicts staining for: (B) nuclei (DAPI, blue), NeuN (red, neuronal marker), dsRNA (magenta) and SARS-CoV-2-S (green); and nuclei (DAPI, blue), ionized calcium-binding adaptor molecule 1 (Iba1, red, microglial marker), dsRNA (magenta) and SARS-CoV-2-S (green). Images were acquired with 630x magnification. Scale bar indicates 50µm. (C) Pearson’s correlation coefficient demonstrating colocalization of SARS-CoV-2-S and dsRNA within GFAP-positive cells.

**Supplementary Figure 4.**
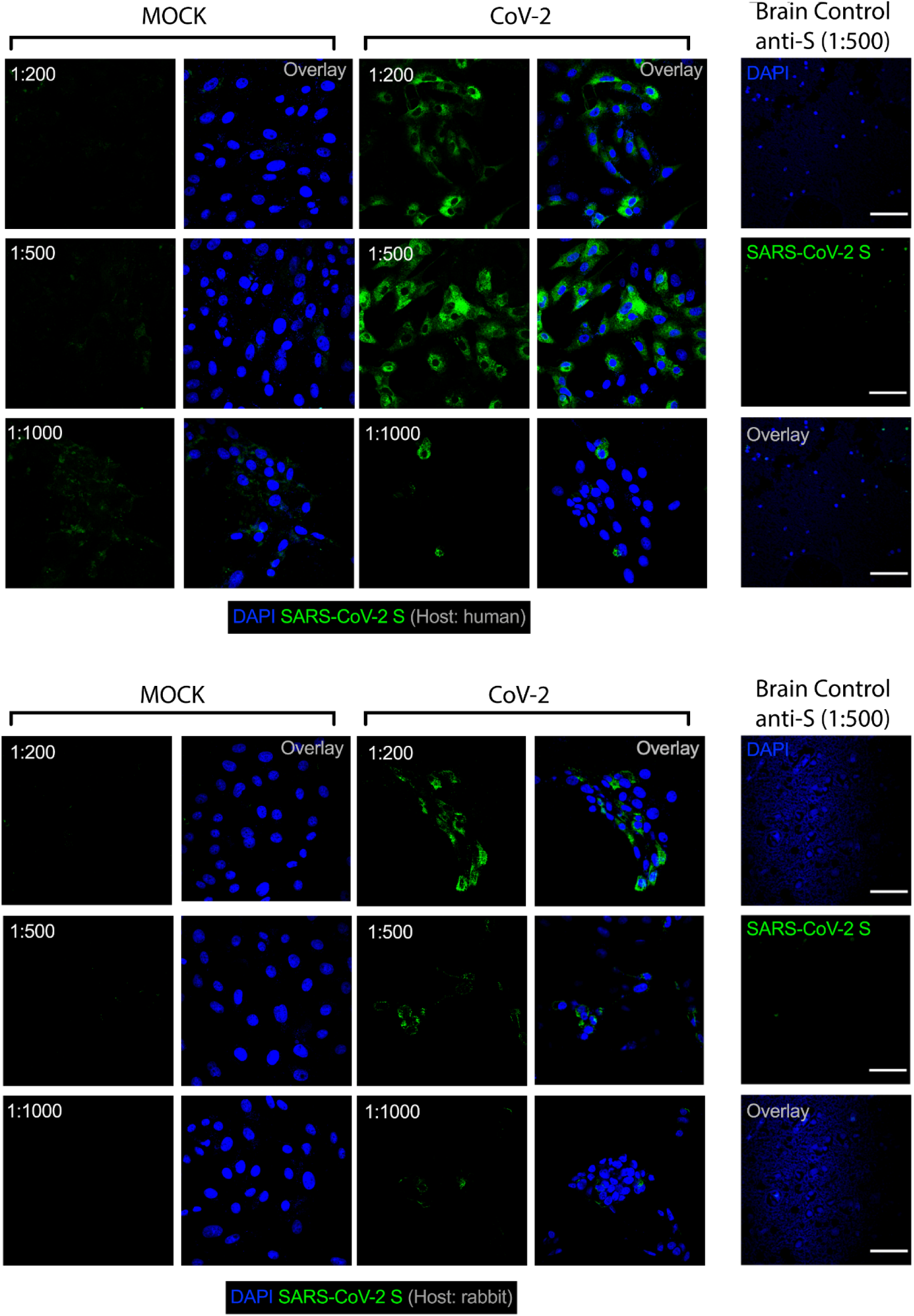
Validation of antibodies against SARS-CoV-2 spike (S) protein. Representative immunostaining and confocal analysis in SARS-CoV-2-infected Vero cells and brain slices from control cases (non-COVID-19) staining with different titrations for anti-S antibodies (1:200, 1:500 and 1:1000). Immunofluorescence images show: nuclei (DAPI, blue) and (A) SARS-CoV-2-S (green, human chimeric monoclonal anti-SARS-CoV-2 spike S1, GeneScript, clone HC2001, Cat. A02038), (B) SARS-CoV-2-S (green, rabbit polyclonal anti-SARS-CoV-2 spike, Rhea Biotech, Cat. IM-0828) or (C) SARS-CoV-2-S (green, rabbit monoclonal anti-SARS-CoV-2 spike, clone T01KHuRb, Cat. 703959). Images were acquired with 630x magnification at the same laser intensity. Scale bar indicates 50µm.

**Supplementary Figure 5.**
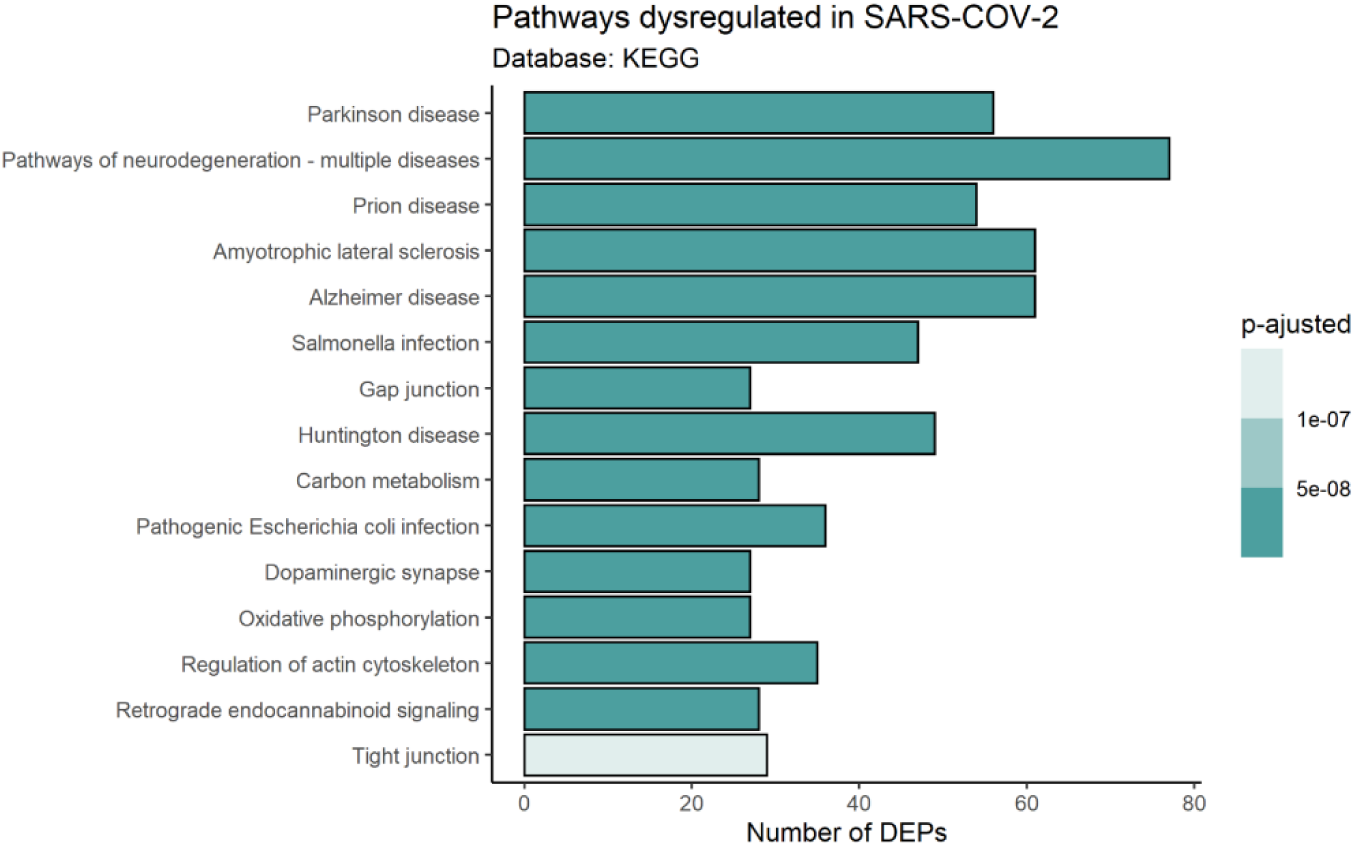
Proteomic analysis of *postmortem* brain tissue from patients who died with COVID-19. Top 15 enriched pathways by differentially expressed proteins in *postmortem* brain tissue from patients who died with COVID-19 (as per KEGG database). Dot color represents lower (blue) or higher (red) expression or no change (gray). Bar color represents the p value adjusted by the false discovery rate (FDR).

**Supplementary Figure 6.**
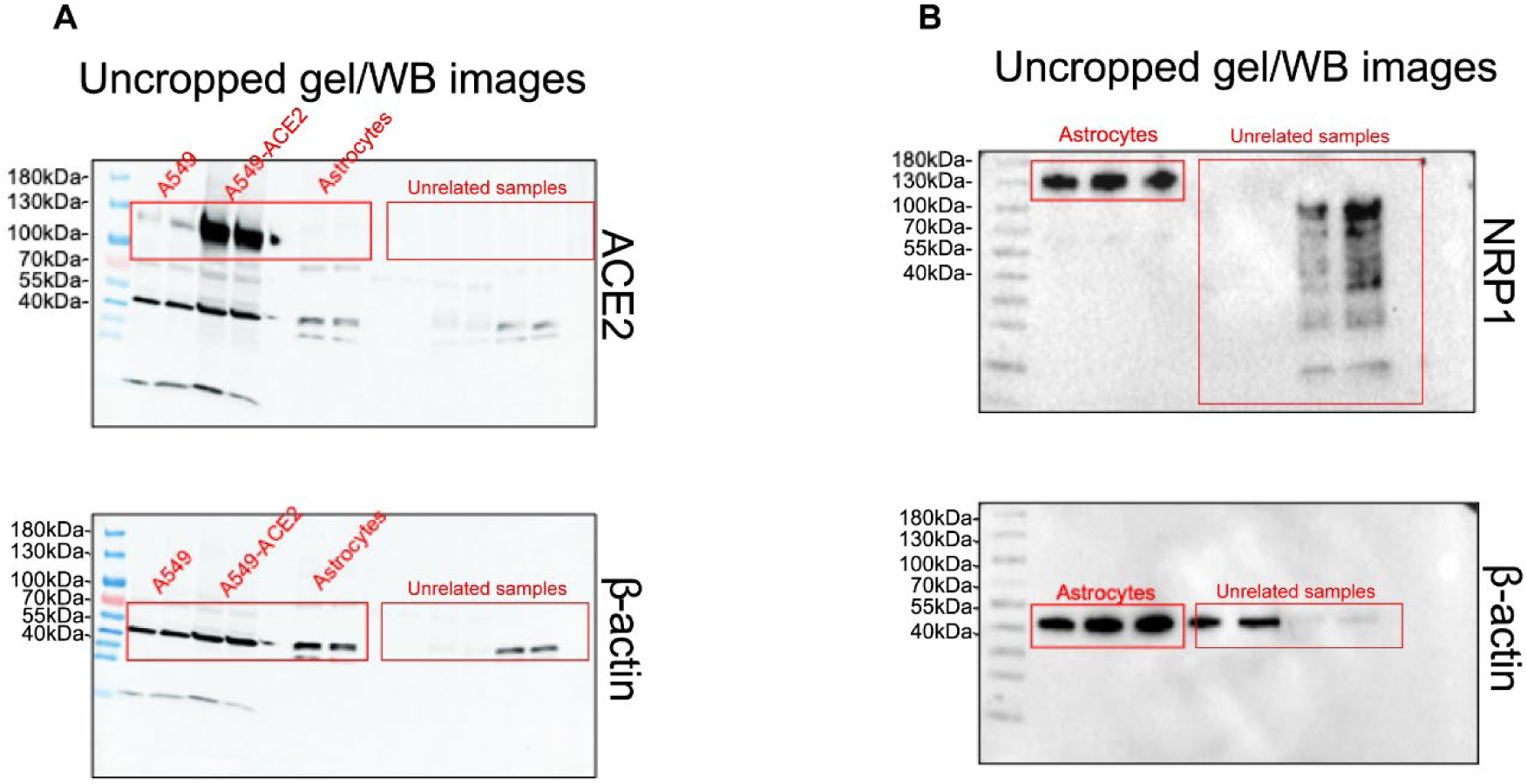
Full scan of the original uncropped immunoblots. (A) Original immunoblots used in Figure 5C for (A) ACE2 and β-actin, and (B) for NRP1 and β-actin. Regions of interest were cropped as indicated in red lines.

**Supplementary Figure 7.**
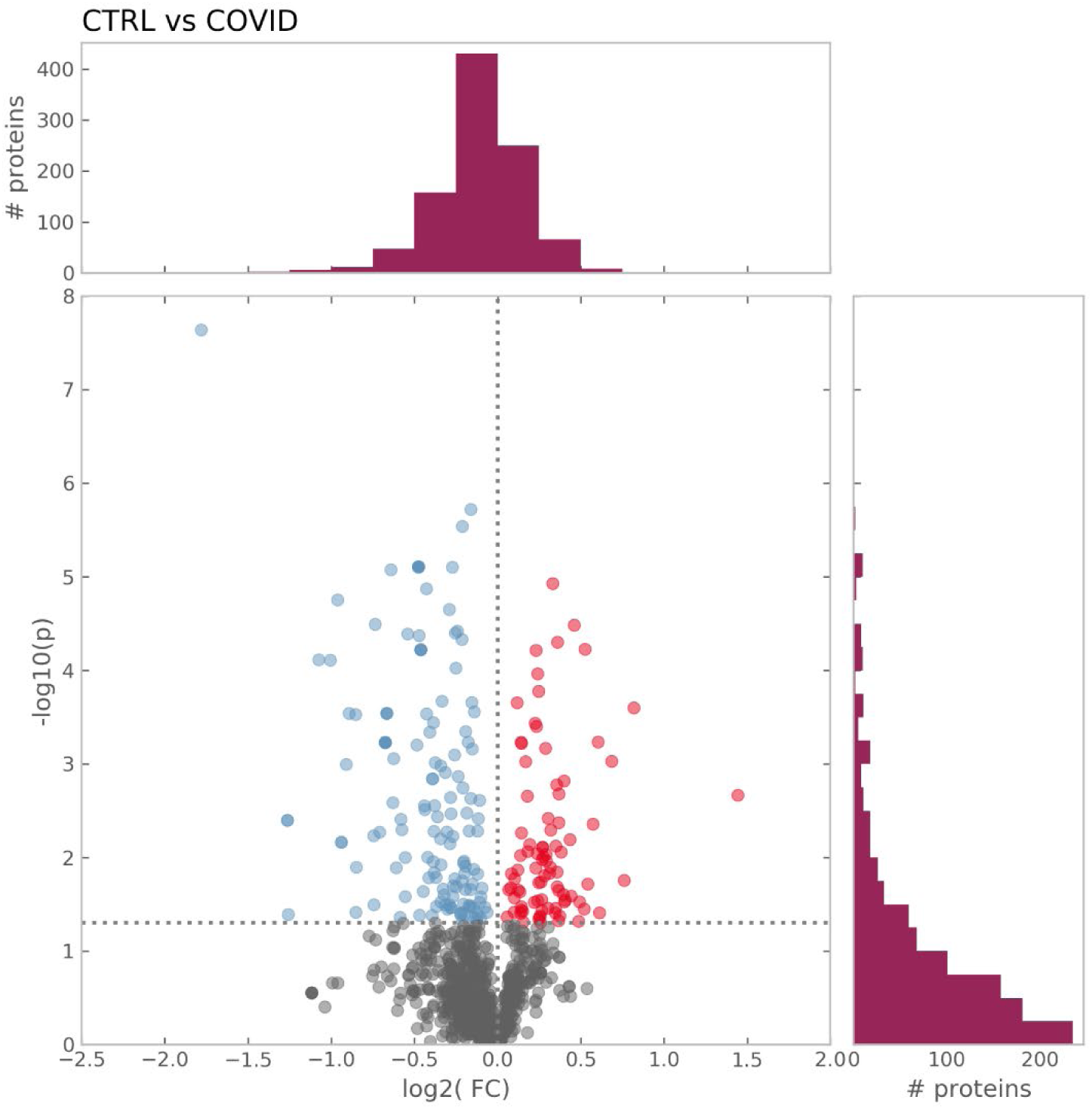
Proteomic analysis of SARS-CoV-2-infected astrocytes. Volcano plot representing all the differentially expressed proteins found in astrocytes after SARS-CoV-2 infection. Dot color represents lower (blue) or higher (red) expression or no change (gray). Bar color represents the p-value adjusted by the false discovery rate (FDR).

**Supplementary Figure 8.**
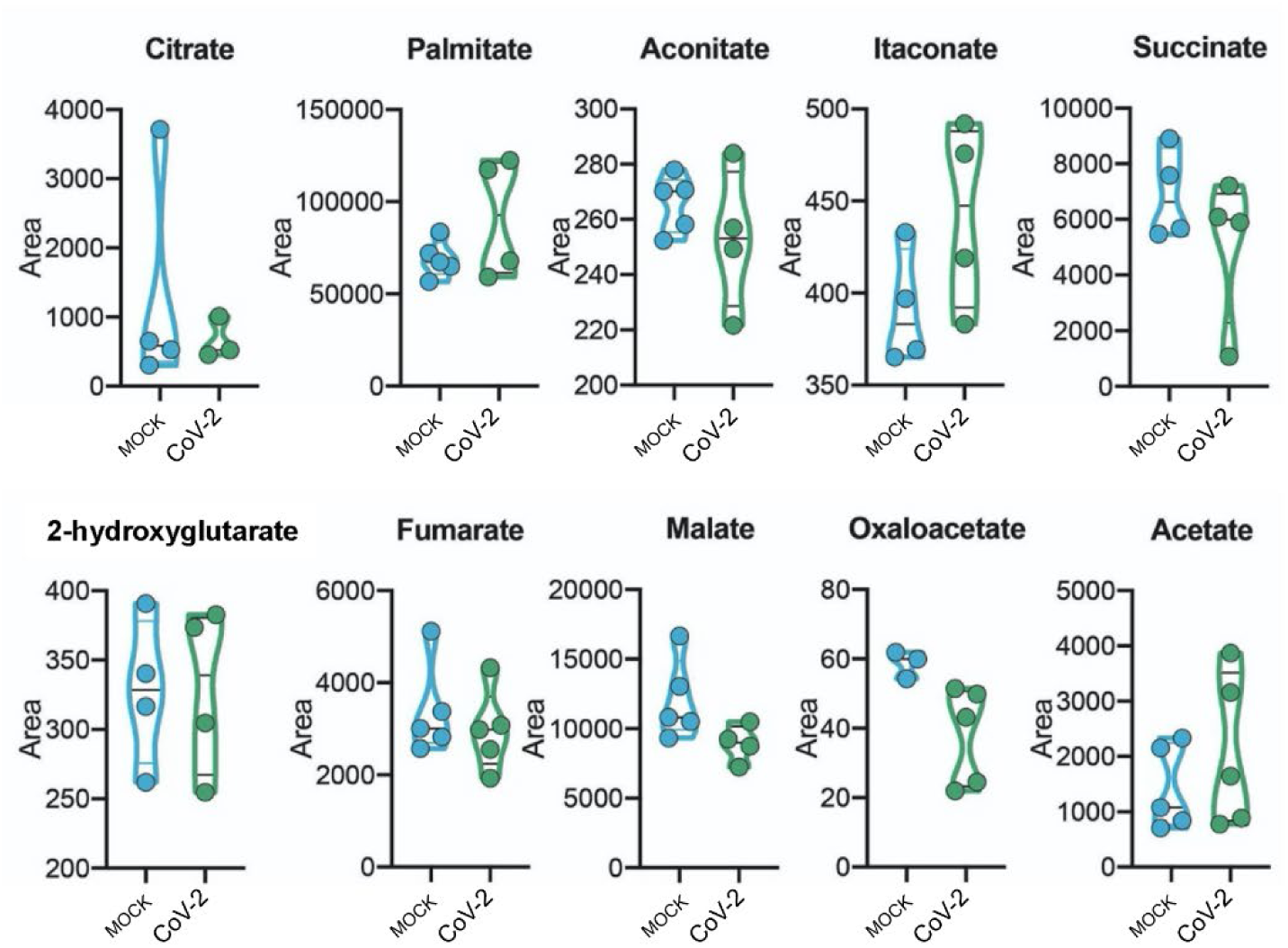
Metabolomic analysis of SARS-CoV-2-infected astrocytes. Human neural stem cell-derived astrocytes were infected *in vitro* with SARS-CoV-2 (MOI 0.1) for 1h, washed thoroughly and harvested after 24h. Mock infection was used as a control. High-resolution mass spectrometry quantification of citrate, palmitate, aconitate, itaconate, succinate, 2-hydroxyglutarate, fumarate, malate, oxaloacetate and acetate in SARS-CoV-2 infected astrocytes *vs.* mock. The integration area of each peak was used to calculate the violin plot graph and an unpaired t-test with Welch’s correction was used for statistical comparison.

**Supplementary Figure 9.**
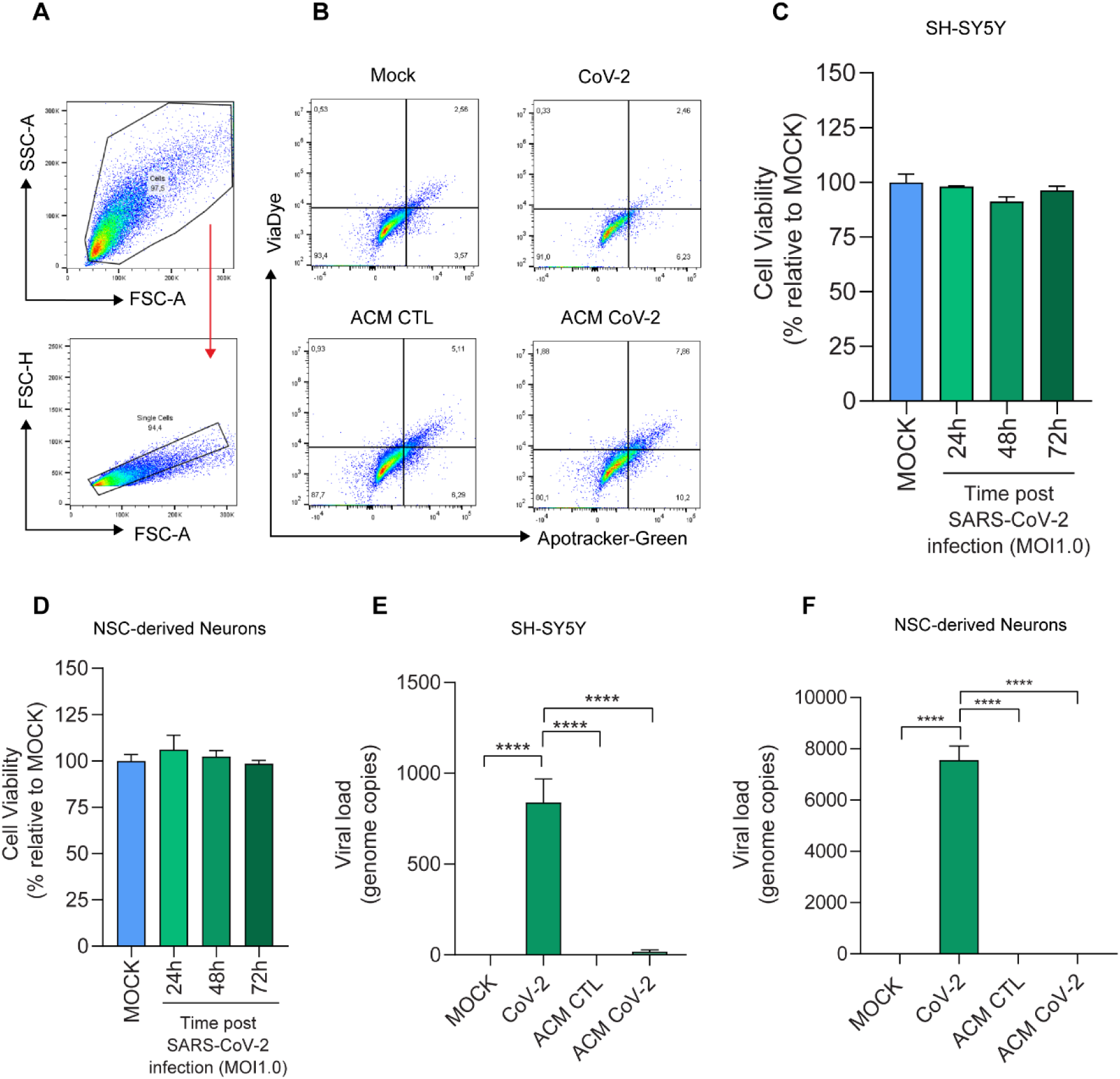
Medium conditioned by SARS-CoV-2-infected astrocytes reduces neuronal viability despite no change from direct virus exposure to neurons. Differentiated SH-SY5Y cells were cultured for 24h in the presence of medium conditioned by SARS-CoV-2-infected astrocytes (ACM CoV-2) or uninfected astrocytes (ACM CTL). Cellular viability was measured by apotracker/fixable viability stain (FVS) and analyzed by flow cytometry. (A) Representative gating strategies. (B) Representative dot-plots of neuronal viability. (C) SH-SY5Y neuronal and (D) NSC-derived neuron cell viability upon SARS-CoV-2 infection. The cells were infected in vitro with SARS-CoV-2 (MOI 1.0) for 1h, washed thoroughly and harvested after 24h. Mock infection was used as a control. SH-SY5Y viability was assessed using calcein AM staining (Invitrogen) at 24, 48 and 72 hours post-infection. The NSC-derived neurons were assessed using an ATP-quantifying, luminescence-based cell viability assay (CellTiter-Glo) at 24h, 48h and 72h post-infection. (E) SARS-CoV-2 viral load detection in differentiated SH-SY5Y neurons and in (F) NSC-derived neurons using RT-PCR after 24h in the presence of mock, SARS-CoV-2 (MOI 1.0) or medium conditioned by either SARS-CoV-2-infected astrocytes (ACM CoV-2) or uninfected astrocytes (ACM CTL). Data are representative of two independent experiments performed in triplicate and shown as mean ± SEM. P-values were determined by one-way ANOVA followed by Tukey’s post hoc test. ***P < 0.001; ****P < 0.0001 compared to the mock group.

**Supplementary Figure 10.**
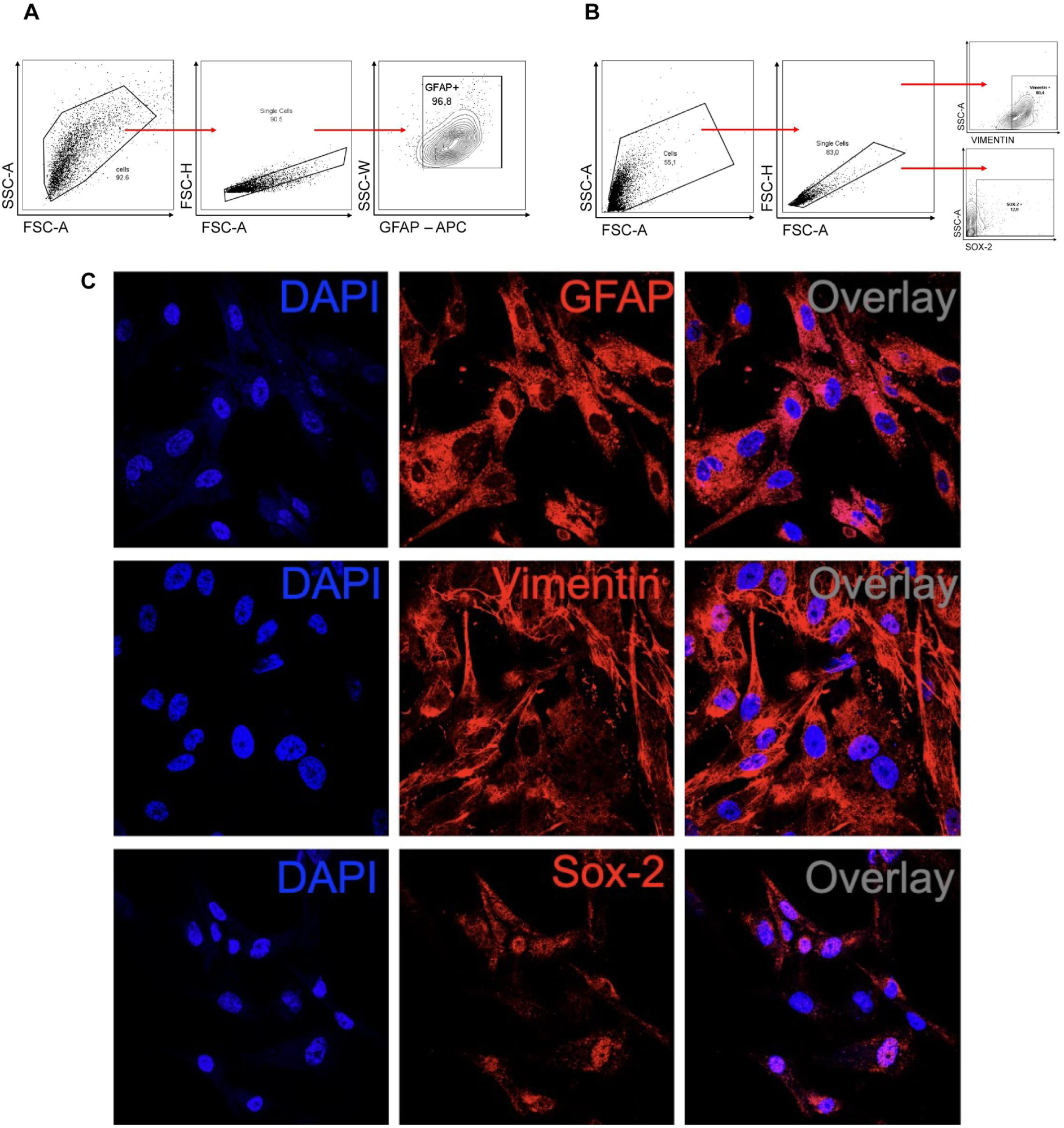
Characterization of human astrocyte cultures. Cells were trypsinized for FACS analysis and incubated with antibodies against GFAP, vimentin and SOX-2. (A) Cells were analyzed and the results were plotted as SSC-A *vs.* GFAP (APC). The percentage of positive cells is indicated in the representative contour plot. (B) Cells were analyzed and the results were plotted as SSC-A *vs.* SOX-2 (APC) and SSC-A *vs.* vimentin (BB515). The percentage of positive cells is indicated in the representative contour plot. (C) Staining for GFAP, vimentin, SOX-2 (red) and nuclei (DAPI, blue). Images were acquired with 630x magnification. Scale bar indicates 50µm.

**Supplementary Figure 11.**
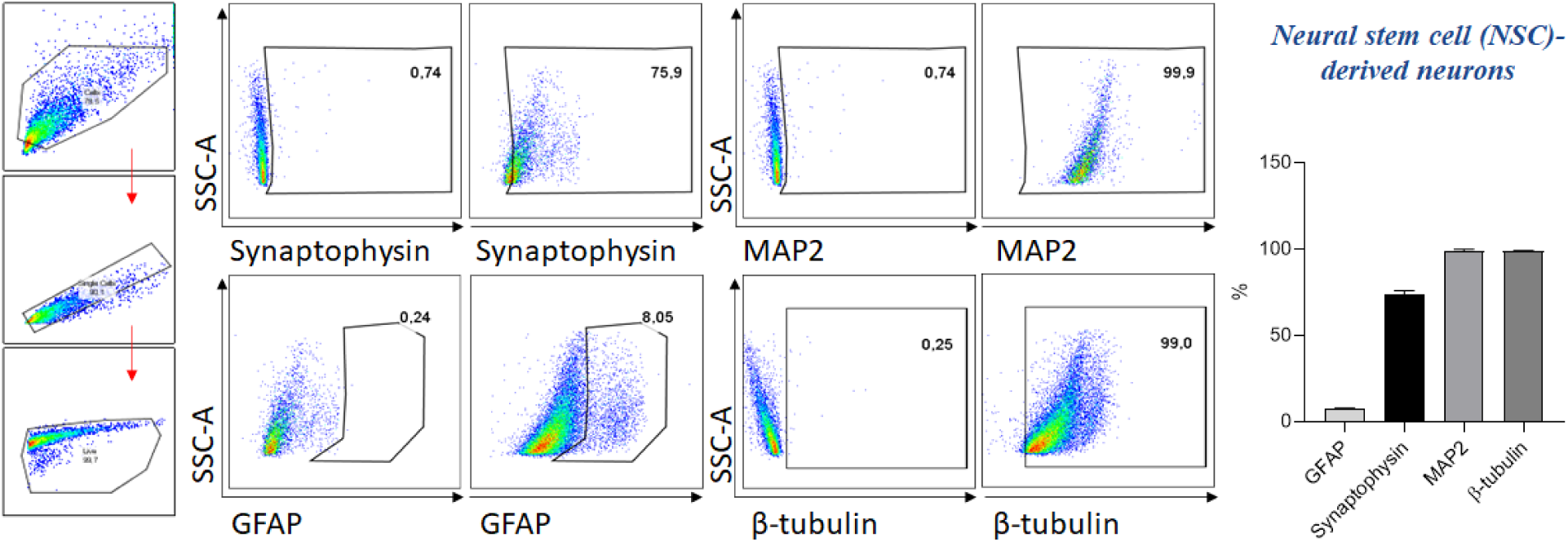
Characterization of human neural stem cell-derived neurons. The cells were trypsinized for FACS analysis and incubated with antibodies against synaptophysin, MAP2, GFAP and β-tubulin. Cells were analyzed and the results were plotted as SSC-A *vs.* synaptophysin, MAP2, GFAP or β-tubulin. The percentage of positive cells is indicated in the representative contour plot.

## Supplementary Table

**Supplementary Table 1.** Reduced cortical thickness in the left hemisphere.

| Value (T statistic) | Cluster size<br>(number of voxels) | Overlap of atlas region | Anatomical correspondence |
| --- | --- | --- | --- |
| 5 | 193 | G_oc-temp_med-Lingual | Lingual gyrus |
| 4.6 | 77 | S_calcarine | Calcarine sulcus |
|  |  | G_cuneus | Cuneus |
|  |  | S_parieto_occipital | Parieto-occipital sulcus |
| 4 | 26 | S_orbital_med-olfact | Olfactory sulcus |
|  |  | G_rectus | Rectus gyrus |

**Supplementary Table 2.** Increased cortical thickness in the right hemisphere.

| Value (T statistic) | Cluster size<br>(number of voxels) | Overlap of atlas region | Anatomical correspondence |
| --- | --- | --- | --- |
| 5.4 | 277 | S_central | Central sulcus |
|  |  | G_precentral | Precentral gyrus |
|  |  | G_postcentral | Postcentral gyrus |
| 4.2 | 40 | G_occipital_sup | Superior occipital gyrus |
|  |  | G_cuneus | Cuneus |

**Supplementary Table 3.** Results of Neuropsychological tests (in z score)

|  | Median | Range |
| --- | --- | --- |
| Logical memory<br>(immediate recall) | -0.63 | -2.25 to 1.96 |
| Logical memory<br>(delayed recall) | -0.27 | -2.37 to 1.58 |
| TRAIL A | -0.7 | -7 to 0.77 |
| TRAIL B | -1.13 | -5.55 to 1.25 |

**Supplementary Table 4.** Partial correlation between BAI scores and cortical thickness of orbitofrontal regions (adjusted for fatigue scores)

|  | Correlation | Significance | FDR adjusted significance |
| --- | --- | --- | --- |
| Left orbital gyrus | -0.19 | 0.2 | 0.4 |
| Right orbital gyrus | -0.42 | 0.004 | 0.016 |
| Left gyrus rectus | -0.017 | 0.91 | 0.91 |
| Right gyrus rectus | -0.1 | 0.5 | 0.67 |

FDR: false discovery rate

**Supplementary Table 5.**
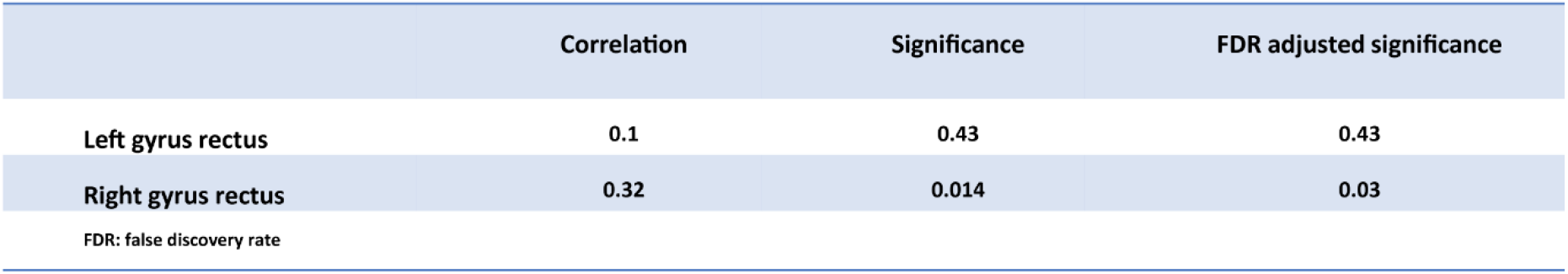
Correlations between TRAIL B scores and thickness of gyrus rectus.

|  | Correlation | Significance | FDR adjusted significance |
| --- | --- | --- | --- |
| Left gyrus rectus | 0.1 | 0.43 | 0.43 |
| Right gyrus rectus | 0.32 | 0.014 | 0.03 |

FDR: false discovery rate

**Supplementary Table 6.** Partial correlations between Logical memory (immediate recall ) scores and cortical thickness of IFG and STG (adjusted for fatigue, depression and anxiety scores)

|  | Correlation | Significance | FDR adjusted significance |
| --- | --- | --- | --- |
| Left IFG (pars opercularis) | 0.27 | 0.08 | 0.1 |
| Right IFG (pars opercularis) | 0.28 | 0.07 | 0.1 |
| Left IFG (pars triangularis) | 0.35 | 0.02 | 0.04 |
| Right IFG (pars triangularis) | 0.43 | 0.004 | 0.02 |
| Left STG (planum temporale) | 0.39 | 0.01 | 0.03 |
| Right STG (planum temporale) | 0.25 | 0.12 | 0.12 |

IFG: inferior frontal gyrus; STG: superior temporal gyrus; FDR: false discovery rate

**Supplementary Table 7:** COVID-19 patient characteristics.

| **Demographics** |  | **%** |
| --- | --- | --- |
| Number | 26 |  |
| Age (years) | 66.08±14.89 |  |
| Hospital day | 12.8±10.08 |  |
| Female | 10 | 38.46% |
| **Comorbidities** |  |  |
| Hypertension | 20 | 76.92% |
| Diabetes | 13 | 50% |
| Obesity | 12 | 46% |
| Lung disease | 2 | 7.69% |
| History of smoking | 8 | 30.76% |
| Heart disease | 10 | 38.46% |
| Kidney disease | 5 | 19.23% |
| Cerebrovascular accident | 4 | 15.38% |
| Cancer | 2 | 7.69% |
| Autoimmune diseases | 3 | 11.53% |
| Immune deficiency | 2 | 7.69% |
| **Laboratory findings** |  |  |
| CRP (mg/dL)* | 13.81±7.125 |  |
| D-Dimers (µg/mL)** | 3.84±2.94 |  |
| LDH (U/L)# | 631.6±481.2 |  |
| Ferritin (ng/mL)& | 1,566±1,381 |  |
| Haemoglobin (g/dL) | 11.73±2.33 |  |
| Neutrophils (cell/mm<sup>3</sup>) | 7,632±4,474 |  |
| Lymphocytes (cell/mm<sup>3</sup>) | 2,955±9,053 |  |
| Platelets (count/mm<sup>3</sup>) | 201,682±122,865 |  |
| **Medications** |  |  |
| Antibiotics | 25 | 96.1% |
| Heparin | 25 | 96.1% |
| Antimalarial | 1 | 3.84% |
| Oseltamivir | 15 | 57.69% |
| Glucocorticoids | 11 | 42.30% |
| **Respiratory status** |  |  |
| Mechanical ventilation | 26 | 100% |
| Nasal-cannula oxygen | 22 | 84.61% |
| pO<sub>2</sub> | 74.45±18.13 |  |
| SatO<sub>2</sub> | 91.80±7.859 |  |
| **Outcome** |  |  |
| Deaths | 26 | 100% |

\*CRP: C-reactive protein (Normal value <0.5 mg/dL);
\*\*D-dimers (NV <0.5 µg/mL); #LDH: lactic dehydrogenase (Normal range: 120-246 U/L); &Ferritin (NR: 10-291 ng/mL)

**Supplementary Table 8:** Clinical data of five COVID-19 patients who manifested histopathological alterations in the brain.

| **Demographics** |  | **%** |
| --- | --- | --- |
| Number | 5 |  |
| Age (years) | 73.0±14.02 |  |
| Hospital day | 7.0±3.317 |  |
| Female | 1 | 20% |
| **Comorbidities** |  |  |
| Hypertension | 5 | 100% |
| Diabetes | 3 | 60% |
| Obesity | 1 | 20% |
| Lung disease | 0 | 0% |
| History of smoking | 3 | 60% |
| Heart disease | 2 | 40% |
| Kidney disease | 1 | 20% |
| Cerebrovascular accident | 0 | 0% |
| Cancer | 0 | 0% |
| Autoimmune diseases | 0 | 0% |
| Immune deficiency | 0 | 0% |
| **Laboratory findings** |  |  |
| CRP (mg/dL)* | 13.91±9.765 |  |
| D-Dimers (µg/mL)** | 6.374±2.526 |  |
| LDH (U/L)# | 1,053.0±878.9 |  |
| Ferritin (ng/mL)& | 2,644±2,057 |  |
| Haemoglobin (g/dL) | 12.72±2.152 |  |
| Neutrophils (cell/mm<sup>3</sup>) | 10,000±6,869 |  |
| Lymphocytes (cell/mm<sup>3</sup>) | 1,780±1,219 |  |
| Platelets (count/mm<sup>3</sup>) | 207,400±122,814 |  |
| **Medications** |  |  |
| Antibiotics | 5 | 100% |
| Heparin | 5 | 100% |
| Antimalarial | 0 | 0% |
| Oseltamivir | 2 | 40% |
| Glucocorticoids | 2 | 40% |
| **Respiratory status** |  |  |
| Mechanical ventilation | 5 | 100% |
| Nasal-cannula oxygen | 5 | 100% |
| Room air | 0 |  |
| pO<sub>2</sub> | 58.98±15.32 |  |
| SatO<sub>2</sub> | 85.78±12.57 |  |
| **Outcome** |  |  |
| Deaths | 5 | 100% |

\*CRP: C-reactive protein (Normal value <0.5 mg/dL);
\*\*D-dimers (NV <0.5 µg/mL); #LDH: lactic dehydrogenase (Normal range: 120-246 U/L); &Ferritin (NR: 10-291 ng/mL)

**Supplementary Table 9:**
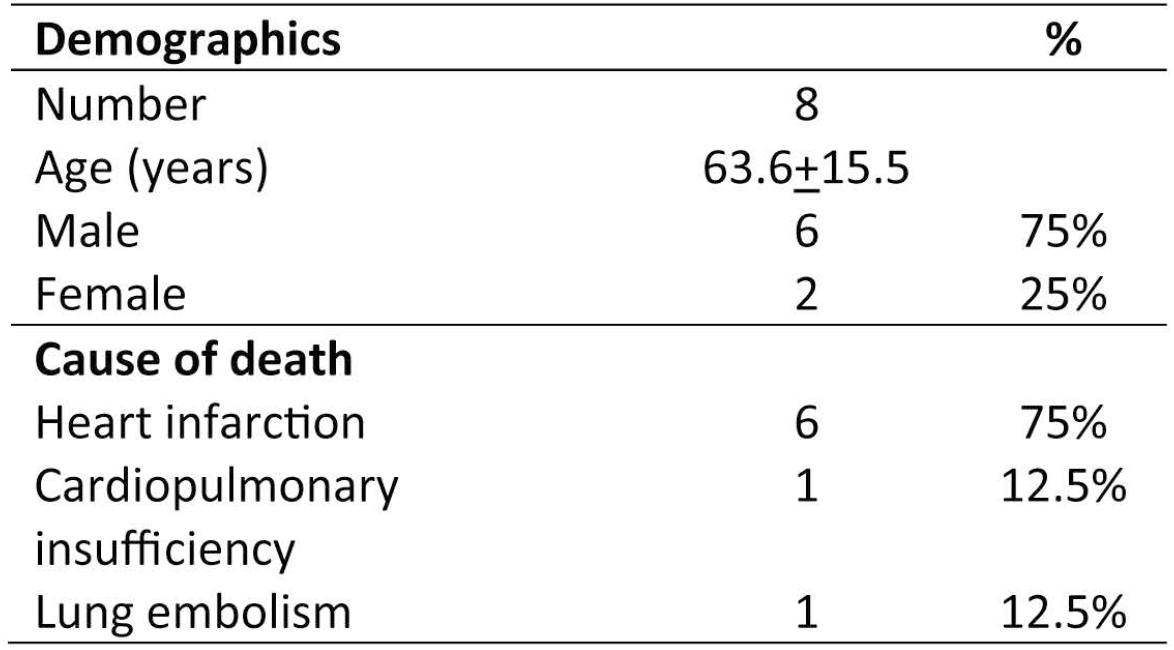
Clinical data of patents controls.

## Supplementary material

### Materials and methods

#### Single-cell transcriptomic analysis

We analyzed single-cell transcriptomic data from the brains of patients with COVID-19 and non-viral controls. The data was generated by Yang et al. ^1^ and made publicly available at https://twc-stanford.shinyapps.io/scRNA_Brain_COVID19. The dataset was downloaded and the RDS file was imported into R environment version v3.6.3. Astrocytes were filtered based on cell type annotations provided by the original authors. To calculate the percentage of cells expressing BSG and NRP1, cell frequency was calculated considering cells with gene count ≥ 1 in the RNA essay for each gene (BSG and NRP1). Cells that met this criterion of minimum expression were considered as expressing the respective gene. A differential expression analysis was conducted using the FindMarkers function in Seurat with the Wilcoxon test comparing COVID-19 astrocyte cells *vs.* non-viral astrocyte cells. More specifically, genes were calculated considering a minimum cellular gene expression of 5% and average log fold change above 0.1. Genes were considered differentially expressed if the adjusted p-value was below 0.05.

#### Immunostaining and confocal microscopy

Brain sections from autopsies and astrocyte cell cultures were fixed with 10% neutral buffered formalin solution or 4% paraformaldehyde (PFA), respectively. Subsequently, the samples were incubated with primary antibodies: mouse monoclonal anti-GFAP Alexa Fluor 488 (EMD Millipore, clone GA5, Cat. MAB3402X, 1:400), human chimeric monoclonal anti-SARS-CoV-2 spike S1 (GeneScript, clone HC2001, Cat. A02038, 1:500), rabbit polyclonal anti-SARS-CoV-2 spike (Rhea Biotech, Cat. IM-0828, 1:200), rabbit monoclonal anti-SARS-CoV-2 Spike (Invitrogen, clone T01KHuRb, Cat. 703959, 1:500), mouse monoclonal anti-double stranded RNA J2 (dsRNA, SCICONS English & Scientific Consulting Kft., clone J2-1909, Cat. 10010200; 1:1,000), rabbit polyclonal anti-IBA1 (FUJIFILM Wako Pure Chemical Corporation, Cat. 019-19741, 1:1,000) and rabbit monoclonal anti-NeuN (Abcam, clone EPR12763, Cat. Ab128886, 1:1,000). The slides were washed twice with TBS-T (Tris-Buffered Saline with Tween 20) and incubated with secondary antibodies donkey anti-mouse IgG AlexaFluor 647 (Thermo Fisher Scientific; Cat. A32787; 1:800) or AlexaFluor 488 (Abcam; Cat. ab150061; 1:800), donkey anti-rabbit IgG AlexaFluor 488 (Abcam; Cat. ab150065; 1:800) or AlexaFluor 594 (Abcam; Cat. ab150076; 1:800) and goat anti-human IgG FITC (Rhea Biotech, Cat. IC-3H04, 1:400). Controls were performed for secondary antibody fluorescence. Antibodies for detecting SARS-CoV-2 in human brain tissue and human astrocytes *in vitro* were first validated with SARS-CoV-2-infected and non-infected Vero cells. In addition, we validated the antibodies in brain sections from controls.

Nuclei were stained with DAPI (Life Technologies; Cat. D1306; 1:1,000). Images were acquired by an Axio Observer combined with an LSM 780 confocal device (Carl Zeiss) with 630x magnification and Z-stack (0.15μm) for brain sections. Colocalization analyses between GFAP and SARS-CoV-2 S1 or GFAP and dsRNA were quantified using Fiji/ImageJ software ^2^. To determine colocalization, we used a ratio of GFAP:SARS-CoV-2 and GFAP:dsRNA in each sample by analyzing Pearson’s correlation coefficients.

#### Western blots

Astrocytic cell culture samples were collected in RIPA Buffer (Sigma Aldrich, Cat. R0278) with protease and phosphatase inhibitors (Cell Signaling, Cat. 5872S). Protein content was quantified using a BCA protein assay kit (Sigma Aldrich, Cat. BCA1). Extracts were separated by 10% SDS-PAGE and transferred to nitrocellulose membranes. After transfering, the membranes were incubated at 4°C with blocking buffer for 2h. The membranes were incubated in 5% BSA solution (Sigma Aldrich, Cat. A7906) and 0.1% Tween 20 (Sigma Aldrich, Cat. P2287) containing rabbit anti-ACE2 polyclonal antibody (Abcam; Cat. ab15348; 1:2,000) or rabbit anti-NRP1 monoclonal antibody (Abcam; clone EPR3113 Cat. ab81321; 1:1,000) overnight at 4°C. Next, the membranes were incubated with anti-rabbit polyclonal antibody (Invitrogen; Cat. 31460; 1:5,000) for 1h. Finally, membranes were incubated with anti-beta-actin (Cell Signaling; clone 8H10D10; Cat. 3700) for 2h at room temperature. Beta-actin expression was used as loading control and was incubated post-ACE2/NRP1 without a stripping process. Fluorescence was detected with an ECL system (Millipore, Cat. WBULS0500) and Chemidoc imaging system (Bio-Rad Laboratories).

#### Human brain slice cultures

Cultures were prepared as previously described ^3,4^. Temporal lobe resections were obtained from two patients who underwent surgery to remove a hippocampal epileptic focus. Collected tissue corresponds to a non-epileptogenic area removed to provide access to the structures to be resected. In our previous works involving adult human brain-derived slice cultures, we have shown the presence of neurons, microglia and astrocytes with no significant morphological/cytoarchitectural alterations, including the preservation of all neuronal cortical layers ^3,4^. Tissue was collected in the surgery room immediately after resection and transported to the laboratory, where it was sliced at 200 µm in a VT1000s automatic vibratome (Leica) and cultivated free-floating with Neurobasal A (Gibco) medium supplemented with 1% Glutamax (Gibco), 1% penicillin/streptomycin (Gibco), 2% B27 (Gibco) and 0.25µg/mL fungizone (Sigma). This procedure was approved by the Ethics Committee of the Ribeirao Preto Medical School (HCRP protocol #17578/15). For viral infection, the medium was removed, and the slices were exposed to a TCID50 of 3×10^6^ for SARS-CoV-2 or an equivalent volume of mock medium. Infection was performed for 2h at 37°C and 5% CO_2_ in a biosafety level 3 laboratory. The inoculum was removed, the tissue was washed and the slices were maintained in fresh medium at 37°C and 5% CO_2_ until processing for subsequent analysis.

#### Proteomics sample preparation, LC-MS/MS analyses and data processing

Astrocytes infected with SARS-CoV-2 and a mock control were collected in biological triplicate. Cells were chemically lysed with lysis buffer (100mM Tris-HCl, 1mM EDTA, 150mM NaCl, 1% Triton-X and protease and phosphatase inhibitors) and mechanically lysed with an ultrasonication probe during 3 cycles of 20s each with 90% frequency on ice. The total protein extract was quantified by BCA, according to the manufacturer’s instructions (Thermo Fisher Scientific, MA, USA). 30µg of total protein extract from each sample was transferred to a Microcon-10 centrifugal filter with a 10kDa cutoff for FASP protein digestion ^5^. Proteins were reduced (10mM DTT) at 56°C for 40min, alkylated by 20min in dark at a room temperature (50mM IAA) and digested overnight by trypsin at 37°C in 50mM ammonium bicarbonate (AmBic), pH 8.0. On the following day, the peptides were recovered from the filter in 50mM AmBic, and trypsin activity was quenched by adding formic acid (FA) to a final concentration of 1% (v/v), whereupon the peptides were desiccated in a SpeedVac and stored at -80°C until use.

Digested peptides were resuspended in 0.1% FA. LC-MS/MS analyses were performed in an ACQUITY UPLC M-Class System (Waters Corporation, Milford, MA) coupled online to a Synapt G2-Si mass spectrometer (Waters Corporation, Milford, MA). 1μg of peptides were loaded onto a trapping column (Symmetry C18 5μm, 180μm × 20mm, Waters Corporation, Milford, MA) and subsequently separated in the analytical column (HSS T3 C18 1.8μm, 75μm × 150mm; Waters Corporation, Milford, MA). For gradient elutions, 0.1% FA was used as solvent A and acetonitrile-FA (99.9% ACN:0.1% FA) as solvent B. A reversed phase gradient was carried out during a 120-minute method, with a linear gradient of 3 - 60% acetonitrile over 90min at 300nL/min. In the Synapt G2-Si, the peptide spectra were acquired by ion mobility-enhanced data-independent acquisition (HDMS^E^). Mass spectrometry analysis was performed in “Resolution Mode”, switching between low (4eV) and high (25–60eV) collision energies, using a scan time of 1.0s per function over 50–2000m/z. The wave velocity for ion mobility separation was 1,000m/s and the transfer wave velocity was 175m/s. A [Glu1]-Fibrinopeptide B standard (Waters Corporation, Milford, MA) was used as the reference lock-mass compound. Each sample was run in three technical replicates.

The raw data from each experiment were processed in Progenesis QI for Proteomics (version 4.0; Waters Corporation, Milford, MA). Tandem mass spectra were searched against the *Homo sapiens* proteome database (UNIPROT, reviewed, release 2020-04), using tolerance parameters of 20ppm for precursor ions and 10ppm for product ions. For peptide identification, carbamidomethylation of cysteines was set as a fixed modification and oxidation of methionines as a variable modification, 2 missed cleavages were permitted and the false discovery rate (FDR) was limited to 1%. Protein identification was performed using a minimum of 1 fragment ion matched per peptide, a minimum of 3 fragment ions per protein and a minimum of 1 peptide per protein.

The label-free quantitative analysis was carried out using relative abundance intensity, normalized by all identified peptides. The expression analysis was performed considering the technical replicates for each experimental condition, following the hypothesis that each group is independent. Proteins with ANOVA (p) ≤ 0.05 between groups were considered differentially expressed.

#### Bioinformatic analyses

Proteomic data visualization was performed in-house in Python (v. 3.7.3). Differentially regulated proteins (p ≤ 0.05) were submitted to systems biology analysis in R (v. 4.0) and Cytoscape environments ^6^. While performing the over-representation analysis (ORA), proteins were enriched using the ClusterProfiler R package ^7^, CellMarker Database ^8^ and Kyoto Encyclopedia of Genes and Genomes (KEGG) ^9^. The network analysis was run as a Reactome Cytoscape plugin ^10^ for module detection and pathway enrichment analysis.

#### Metabolomics sample preparation, UPLC-MS/MS analyses and data processing

Astrocytes were washed twice with PBS at physiological pH, then the cells were collected with 600μL of methanol. Samples were dried and stored at -80°C until the metabolite extraction step. 473μL of water, 600μL of methanol and 1168μL of chloroform were added and the tubes were shaken vigorously for 2 minutes. Subsequently, samples were centrifuged for 5min at 13,000 x*g*. The aqueous supernatant and the organic phase (lower phase) were collected and dried for 60min and 40min, respectively, in a vacuum concentrator. All samples were stored at -80°C until analysis by LC-MS/MS.

The samples were resuspended in 100μL of a methanol:water mixture (1:1) and for each analysis, 4μL of the sample was injected. Sample separation was performed by hydrophobic interaction liquid chromatography (HILIC) using an Acquity UPLC® BEH amide column (1.7µm, 2.1mm x 100mm). The mobile phases used for the separations were ACN:H_2_O (80:20) as mobile phase A and ACN:H_2_O (30:70) as mobile phase B, both phases containing 10mM ammonium formate and 0.1% ammonium hydroxide. Separation was then performed by a gradient from 99 - 1% buffer A over 7min. The column was returned to 99% buffer A for 2min for re-equilibration before the next injection for a total run time of 10min. Data acquisition was performed in negative mode and the instrument was operated in MS^E^ mode in the m/z range of 50–800Da, with an acquisition time of 0.1s per scan.

Identification of the metabolites of interest was carried out manually by spectral features, and the level 3 identification was obtained according to Schrimpe-Rutledge et al. ^11^ using 5ppm as the error cutoff. The integration area of each peak was used to calculate the violin plot and an unpaired t-test with Welch’s correction was used for statistical comparison. All analyses were performed using GraphPad Prism 8.0 software (San Diego, CA, USA) and a significance level of p ≤ 0.05 was adopted.

#### RNA extraction, gene expression analyses and viral load

Total RNA extraction was performed using TRI Reagent according to the manufacturer’s instructions (Sigma, St Louis, USA). RNA concentration was determined by a DeNovix spectrophotometer and RNA integrity was assessed by visualization of 28S and 18S ribosomal RNA on a 1% agarose gel. Reverse transcription was performed with 0.5µg of RNA using a GoScript reverse transcriptase kit (Promega, Madison, WI, USA) according to the manufacturer’s instructions. qPCR was performed using astrocyte cDNA diluted 1:10 and the qPCR SybrGreen Supermix (Qiagen, Valencia, CA, USA) containing forward and reverse primers in RNAse-free water. All reactions were performed in a CFX384 Touch real-time PCR detection system (Biorad, Hercules, CA, USA) and cycling conditions were set as follows: 50°C for 2min; 95°C for 10min; (95°C for 15s; 60°C for 1min) x 40 cycles. To evaluate primer specificity, a melting curve analysis was performed by heating samples from 65°C to 99°C (1°C increment changes at 5s intervals). All sample measurements were performed in duplicate. Primers were designed with PrimerBlast and used at a concentration of 200nM. Data were normalized to the expression of 18S (Fwd 5’ CCCAACTTCTTAGAGGGACAAG 3’; Rev 5’ CATCTAAGGGCATCACAGACC 3’) and the relative quantification value of each target gene was determined using a comparative CT method^12^. For virus detection, SARS-CoV-2 nucleocapsid N1 primers were used as previously described (Fwd 5’ CAATGCTGCAATCGTGCTAC 3’; Rev 5’ GTTGCGACTACGTGATGAGG 3’) 13,14.Data were expressed as mean ± SEM. Statistical significance was calculated by a two-tailed unpaired Student’s t-test. All analyses were performed using GraphPad Prism 8.0 (San Diego, CA, USA) and a significance level of p ≤ 0.05 was adopted.

#### Astrocyte bioenergetics

Astrocytes were plated on Seahorse XF-24 plates at a density of 1.5×10^4^ cells per well and incubated in complete culture medium for two days at 37°C in 5% CO_2_. 24 hours before the experiment, cells were either infected by SARS-CoV-2 (MOI 0.1) or not infected (MOCK). One day post-infection, the culture medium was changed to Seahorse Base medium (supplemented with 1mM pyruvate, 2mM glutamine and 10mM glucose) and cells were incubated at 37°C in a non-CO_2_ incubator for 1h. OCR (Oxygen Consumption Rate) was measured over the course of the experiment under basal conditions and after injections of oligomycin (1µM), FCCP (5µM) and antimycin A/rotenone (100nM/1µM). Protein concentration was determined for each well to normalize the data. Data were expressed as mean ± SEM of two independent experiments performed in quintuplicate. Statistical significance was calculated by two-tailed unpaired Student’s t-test. All analyses were performed using GraphPad Prism 8.0 software (San Diego, CA, USA) and a significance level of p ≤ 0.05 was adopted.

#### Differentiation of the SH-SY5Y human neuroblastoma cell line

The SH-SY5Y cell line (SH-SY5Y-ATCC-CRL-2266) was cultivated using a commonly used neuronal differentiation protocol ^15–17^ using DMEM/F12 medium, 10% FBS and 1% penicillin-streptomycin at 37°C in humidified air with 5% CO_2_. The SH-SY5Y cells were plated and, upon reaching 25-30% confluency, the medium was changed to neuronal differentiation medium consisting of DMEM/F12 with 1% FBS and 10µM retinoic acid (Sigma Aldrich). The differentiation medium was replaced every 2-3 days during 2 weeks. These differentiated SH-SY5Y cells are more closely related to adrenergic neurons, but they also express dopaminergic markers^17^. The SH-SY5Y cell line was kindly donated by Prof. Dr. Gustavo J. S. Pereira (Unifesp).

#### NSC differentiation into neurons

The human NSC-derived neurons were cultivated following the protocol described by Thermo Fisher Scientific ^18–20^. NSCs were plated on geltrex-coated plates and maintained with NEM medium at 37°C in humidified air with 5% CO_2_. Upon reaching 40% confluency, the medium was changed to neuronal differentiation medium consisting of DMEM/F12 and Neurobasal medium (1:1) with 1% B27 supplement (Thermo Fisher Scientific, Carlsbad, CA, USA) and 1% Glutamax (Thermo Fisher Scientific, Carlsbad, CA, USA). The medium was renewed every four days for 20 days by removing half of the volume and adding the same volume of fresh medium. Medium renewal was performed in this manner since factors secreted by the differentiating cells are important for successful differentiation. Two control cell lines were used: GM23279A, obtained from a female subject (available at Coriell) and BR-1 ^21^. Both cell lines were cultivated following the protocol described by Thermo Fisher Scientific ^22–25^.

These cells and protocols have been extensively characterized elsewhere ^18–20^. We also used FACS analysis for cellular markers and found a bonafide neuronal phenotype due to the expression of the neuronal markers synaptophysin (75.9% of cells), MAP2 (99.9%) and β-tubulin (99%) and astrocytic marker GFAP (8.1%) (Supplementary Fig. 11).

#### CellTiter-Glo luminescent cell viability assay

Astrocyte and NSC-derived neuron death caused by SARS-CoV-2 infection was measured using ATP quantification by a luminescence assay (Promega, Madison, WI, USA; G7572). The CellTiter-Glo luminescent cell viability assay determines the number of viable cells in culture through quantitation of ATP levels, which reflect the presence of metabolically active cells. Astrocytes and NSC-derived neurons were infected with SARS-CoV-2 and harvested after 24, 48 or 72h. Astrocytes and NSC-derived neurons were then washed and the CellTiter-Glo reagent was added to the cells, following the manufacturer’s instructions (Promega; G7572). The luminescent signals were obtained using a FlexStation 3 (Molecular Devices, CA, USA).

#### Calcein AM cell viability assay

SH-SY5Y death caused by SARS-CoV-2 infection was measured using Calcein AM, a cell-permeant dye that can be used to determine cell viability in most eukaryotic cells. In live cells, the non-fluorescent calcein AM is converted to green, fluorescent calcein, after acetoxymethyl ester hydrolysis by intracellular esterases (Invitrogen, Cat. No. C3100). The SH-SY5Y cell line was infected with SARS-CoV-2 and harvested after 24, 48 or 72h. SH-SY5Y cells were then washed and the approximately 2µM serum-free calcein AM working solution (FluoroBrite™ DMEM, ThermoFisher Scientific) was added to the cells and incubated for 30 minutes at room temperature, according to the manufacturer’s protocol. The working solution was removed and replaced with a normal growth medium. The fluorescence signals were obtained using a GloMax® Discover microplate reader (Promega, Madison, WI, USA).

#### Exposure of neurons to astrocyte-conditioned media

Astrocytes, human NSC-derived neurons and SH-SY5Y cells were cultured separately in standard conditions until complete differentiation. First, astrocytes were infected with SARS-CoV-2 (MOI 0.1 and MOI 1.0) or mock condition (MOCK), and after 24 hours, the medium was removed and cells were washed with PBS and cultured for 24 hours. The media of NSC-derived or SH-SY5Y neurons were removed and replaced by the conditioned media from SARS-CoV-2-infected astrocytes or control medium for 24 hours. After incubation, cells and their medium were collected for flow cytometry analysis, following the procedure described below. We also evaluated the cell viability of SH-SY5Y cells exposed to astrocyte-conditioned media using CellTiter-Glo luminescent cell viability assay.

#### Flow cytometry analyses

The expression of GFAP, synaptophysin, MAP2 and β-tubulin was evaluated through FACS analysis. Astrocytes and NSC-derived neurons were collected and stained with BD Horizon fixable viability stain 510 for 30min at 4°C. Primary antibodies anti-GFAP (Abcam; Cat. Ab7260); anti-synaptophysin D35E4 (Cell Signaling; Cat.#5461); XP Rabbit mAb (Cell Signaling; Cat. #5461); anti-MAP2 (Abcam; Cat. ab32454); and anti-tubulin, beta III isoform, C-terminus (Milipore; Cat. #MAB1637) all diluted in BD Perm/Wash buffer, were added to the cells and incubated for 1h at 4°C (1:500). Secondary antibodies donkey anti-mouse IgG AlexaFluor 594 (Cell Signalling; Cat. 8890S), donkey anti-rabbit IgG AlexaFluor 647 (Abcam; Cat. ab15006) and donkey anti-goat IgG AlexaFluor 488 (Abcam; Cat. ab150129), all diluted in BD Perm/Wash buffer, were added and incubated for 30min at 4°C (1:250). Cells were washed with BD Perm/Wash buffer and then transferred to polypropylene FACS tubes. Analyses were carried out on a FACSymphony (Becton & Dickinson, San Diego, CA, USA).

The viability of human NSC-derived and SH-SY5Y neurons was determined 24h after incubation with the conditioned medium of SARS-CoV-2-infected astrocytes or control medium. The percentage of live (Apotracker-/FVS510-), necrotic (Apotracker-/FVS510+), early apoptotic (Apotracker+/FVS510-) and late apoptotic (Apotracker+/FVS510+) cells was determined by flow cytometry (FACSymphony, Becton & Dickinson, San Diego, CA, USA), after labeling with fixable viability stain (FVS510, BD Biosciences, #564406) and Apotracker Green (BioLegend, #427403). Data were analyzed using FlowJo software (BD Biosciences). Data are representative of at least two independent experiments performed in triplicate and are shown as mean ± SEM. P values were determined by one-way ANOVA followed by Tukey’s post hoc test.

### Data availability

The mass spectrometry proteomic data have been deposited to the ProteomeXchange Consortium via the PRIDE ^26^ partner repository with the dataset identifiers PXD023781 and 10.6019/PXD023781.

## ACKNOWLEDGMENTS

We thank Edison Durigon for providing the SARS-CoV-2 virus. We thank Gabriela Lopes Vitória, Elzira E. Saviani and Paulo Baldasso for technical support. We dedicate this work to Dr. Nilton Barretos dos Santos, a brilliant scientist who was one of the many young victims of COVID-19.

